# Minimum concurrent sleep, physical activity, and nutrition variations associated with lifeSPAN and healthSPAN improvements: a population cohort study

**DOI:** 10.1101/2025.04.24.25326344

**Authors:** Nicholas A. Koemel, Raaj K. Biswas, Matthew N. Ahmadi, Armando Teixeira-Pinto, Mark Hamer, Leandro F. M. Rezende, John Mitchell, Rebecca M. Leech, Mouna Sawan, Margaret Allman-Farinelli, Dorothea Dumuid, Adrian Bauman, Carol Maher, Stephen Barrett, Clara Chow, Alice A. Gibson, David Raubenheimer, Samantha L. Hocking, Kathryn Williams, Peter A. Cistulli, Stephen J. Simpson, Emmanuel Stamatakis

**Affiliations:** Mackenzie Wearables Research Hub, Charles Perkins Centre, University of Sydney, Sydney, NSW, Australia; School of Health Sciences, Faculty of Medicine and Health, University of Sydney, Sydney, NSW, Australia; Charles Perkins Centre, University of Sydney, Sydney, NSW, Australia; Centre for Precision Data Science, University of Sydney, Sydney, NSW, Australia; School of Public Health, Faculty of Medicine and Health, University of Sydney, Sydney, NSW, Australia; Institute Sport Exercise and Health, Division of Surgery and Interventional Science, University College London, United Kingdom; National Institute for Health Research, University College London, London, United Kingdom; Department of Preventive Medicine, Escola Paulista de Medicina, Universidade Federal de São Paulo, São Paulo, Brazil; Chronic Disease Epidemiology Research Center, Universidade Federal de São Paulo, São Paulo, Brazil; Faculty of Health Sciences, Universidad Autónoma de Chile, Providencia 7500912, Chile; Institute for Physical Activity and Nutrition (IPAN), School of Exercise and Nutrition Sciences, Deakin, Geelong, VIC, Australia; School of Pharmacy, Faculty of Medicine and Health, University of Sydney, Sydney, NSW, Australia; School of Nursing, Faculty of Medicine and Health, University of Sydney, Sydney, NSW, Australia; Alliance for Research in Exercise, Nutrition and Activity, Allied Health and Human Performance, University of South Australia, Adelaide, SA, Australia; Bendigo Health and Holsworth Research Initiative, La Trobe Rural Health School, Bendigo, VIC, Australia; Westmead Applied Research Centre, Faculty of Medicine and Health, University of Sydney, Sydney, Australia; Leeder Centre for Health Policy, Economics and Data, Sydney School of Public Health, Faculty of Medicine and Health, University of Sydney, Sydney, NSW, Australia; School of Life and Environmental Sciences, Faculty of Science, University of Sydney, Sydney, NSW, Australia; Metabolism and Obesity Service, Royal Prince Alfred Hospital, Sydney, NSW, Australia; Central Clinical School, Faculty of Medicine and Health, University of Sydney, Sydney, Australia; Department of Endocrinology, Nepean Hospital, Kingswood, NSW, Sydney, Australia; Department of Respiratory and Sleep Medicine, Royal North Shore Hospital, Sydney, NSW, Australia

**Keywords:** Healthspan, lifespan, sleep, nutrition, diet, physical activity, exercise, lifestyle risk factors, life expectancy, disease free life expectancy

## Abstract

**Background:** Sleep, physical activity, and nutrition (SPAN) are key determinants of both life expectancy (lifespan) and disease-free life expectancy (healthspan) yet are often studied and promoted in isolation. This study examined the joint association of these three behaviours with lifespan and healthspan.

**Methods:** Prospective cohort analysis of 59,078 participants from the UK Biobank accelerometry sub-study (median age: 64.0 years; 45.4% male) who wore accelerometers for 7 days and self-reported dietary data. Moderate to vigorous physical activity (MVPA; mins/day) and sleep (hours/day) were calculated using a validated wearables-based algorithm. Diet quality was assessed using a 10-item diet quality score (DQS) based on the consumption of vegetables, fruits, whole grains and refined grains, unprocessed and processed meats, fish, dairy, vegetable oils, and sugary beverages (ranging 0-100, where higher values indicate a higher diet quality). Lifespan and healthspan (defined as years lived free of cardiovascular disease (CVD), cancer, type 2 diabetes (T2D), chronic obstructive pulmonary disease (COPD), and dementia) were estimated across 27 joint tertile combinations of SPAN behaviours and a continuous composite SPAN score using a life table approach. Mortality rates in the life table were adjusted using hazard ratios from a Cox proportional hazards model. The minimum meaningful dose was defined as the smallest combination of behavioural changes associated with a statistically significant gain in lifespan or healthspan.

**Results:** During a median follow-up of 8.1 years, 2,458 deaths, 9,996 incident CVD, 7,681 cancers, 2,971 T2D, 1,540 COPD, and 508 dementia events occurred. Compared with participants in the least favourable tertiles for all SPAN behaviours, those in the optimal tertiles (7.2-8.0 hours/day of sleep; >42 mins/day of MVPA; a DQS of 57.5-72.5) had 9.35 additional years of lifespan (95% CI: 6.67, 11.63) and 9.45 additional years of healthspan (95% CI: 5.45, 13.61). When compared to the 5^th^ percentile for all SPAN behaviours, a minimum combined improvement of 5 mins/day of sleep, 1.9 mins/day MVPA, an extra 5-point increase in DQS (e.g., an additional ½ serving of vegetables/day) was associated with 1 additional year of lifespan (95% CI: 0.77, 1.26). For healthspan, a combined improvement of 18.6 mins/day of sleep, 3.4 mins/day of MVPA, and a 21-point diet score increase (e.g. additional 1 cup of vegetables and two servings per week of fish) was associated with 4.0 additional disease-free years (95% CI: 0.27, 8.11).

**Conclusion:** Small, concurrent improvements in sleep, physical activity, and diet quality were associated with substantial gains in lifespan and healthspan. These findings highlight a pragmatic and synergistic approach to improving population health through modest, combined behavioural changes.

## INTRODUCTION

Sleep, physical activity, and nutrition (SPAN) are key modifiable risk factors for noncommunicable diseases (NCD) and all-cause mortality^1-3^. Both insufficient and excessive sleep have been shown to disrupt normal metabolic and brain health through mechanisms such as impaired glycaemic control, inflammation, and dysregulation of appetite hormones^4,5^. Physical inactivity contributes to one in six deaths in the UK^6^ and plays a major role in the progression of age-related chronic diseases^7^. Poor diet quality and excess caloric intake adversely affect cardiometabolic health and are central to the global obesity epidemic^8,9^.

Collectively, SPAN behaviours are linked with increased risk of leading causes of NCD-related morbidity and mortality, including cardiovascular disease (CVD)^10^, certain cancers^3^, Type 2 diabetes^11^, dementia^12,13^, and respiratory conditions such as chronic obstructive pulmonary disease (COPD)^14^. Despite increases in average the life expectancy (lifespan) over recent decades^15^, healthspan (defined as years lived free of major chronic disease)^16^ has remained stagnant or declined^17^. In the UK, it is projected that by 2035, over two-thirds of adults aged ≥35 years will be living with multiple chronic conditions^18^, likely resulting in greater healthcare expenditure and reduced quality of life^19^. Modifying SPAN behaviours presents a viable strategy to extend healthspan by influencing the trajectory of biological aging^20^ and delaying the onset of chronic disease^21^. However, these behaviours are often examined independently, neglecting the complex behavioural and physiological interdependencies between them^5,22,23^. For instance, inadequate sleep duration may impair appetite regulation, leading to higher energy intake^5^, while inadequate diet quality may reduce sleep quality. Similarly, sleep deprivation may increase fatigue and reduce physical activity^5,24,25^. Emerging evidence supports a synergistic relationship among SPAN behaviours. A recent prospective study of over 59,000 UK adults^26^ found that modest concurrent improvements in all three behaviours were associated with meaningful reductions in mortality risk. Specifically, a theoretical minimum combined increase of just 15 mins/day of sleep, 1.6 mins/day of moderate-to-vigorous physical activity (MVPA), and a 5-point increase in diet quality score (e.g., an additional ½ serving of vegetables or one fewer serving of processed meat per week) was associated with a 10% reduction in all-cause mortality risk. In isolation, achieving this level of risk reduction required 60% more sleep (additional 24 mins/day), 25% more MVPA (2 mins/day), while diet alone was unable to achieve this level of risk reduction.

Most prior studies examining lifestyle behaviours in relation to life expectancy^27-31^ and healthspan^32-35^ have relied on broad, self-reported behavioural categories (e.g., active vs. inactive), limiting granularity and interpretability. To our knowledge, no study has examined the combined associations of SPAN behaviours with lifespan and healthspan using high-resolution, interpretable metrics (e.g., minutes per day of activity; servings of fruits per day). In this study, we applied a multidimensional behavioural framework^26^ to examine the combined associations of device-measured sleep and physical activity, along with a detailed diet quality score, with life expectancy and chronic healthspan^36^. Our primary goal was to estimate the minimum theoretical behavioural improvements associated with statistically meaningful gains in lifespan and healthspan.

## METHODS

### Study Population

We used data from the UK Biobank accelerometry sub-study^37^, part of a larger cohort comprising 502,629 adults aged 40-69 recruited from 2006 to 2010. Participants completed touchscreen questionnaires for sociodemographic, lifestyle characteristics, and health status at recruitment. All participants provided written informed consent. Ethical approval was granted by the UK National Health Service (NHS) and National Research Ethics Service for the UK (No. 11/NW/0382).

### Outcome Ascertainment

Participants were followed through November 30^th^, 2022. Data on all-cause mortality were retrieved via linkage from the National Health Service Central Register and the National Records of Scotland. Incidence of five major noncommunicable diseases (CVD, cancer, type 2 diabetes, COPD, and dementia) was ascertained using inpatient hospital records. Additional cancer data linkage was obtained through national cancer registries. A complete list of ICD-10 codes and outcome definitions is provided in **Supplementary Methods 1**.

### Sleep, Physical Activity, and Nutrition Assessment

Between 2013 and 2015, a subsample of 103,684 participants was invited to wear wrist-worn accelerometers (Axivity AX3, York, UK) on their dominant wrist for 7 days^37^. As previously described^38-43^, participants were included in the analysis if they had valid data for at least three days (>16 hours/day), including at least one weekend day. We excluded participants with no sleep data, faulty devices, poor calibration, self-reported inability to walk, or incomplete covariate data (**Supplemental Figure 1**).

Sleep duration (hours/day) was estimated using a validated algorithm based on wrist tilt angle variation in 5-second epochs^12,44,45^. Moderate-to-vigorous physical activity (MVPA; mins/day) was calculated using a validated two-stage machine learning algorithm that classifies activity type, then intensity, in 10-second epochs^38,39,42,46,47^. Additional details regarding the development, validation and performance of this schema are provided in **Supplementary Methods 2**.

Diet quality was assessed using a 29 item food-frequency questionnaire (FFQ) completed at recruitment (2006-2010) covering consumption of commonly eaten foods over the past 12 months^48^. A validated Diet Quality Score (DQS)^26,49^ was computed, ranging from 0 (poorest diet) to 100 (healthiest diet), based on intake of vegetables, fruits, fish, dairy, whole grains, and vegetable oils and lower intake of refined grains, processed meats, unprocessed red meats, and sugar-sweetened beverages (**Supplemental Table 1**).

### Statistical Analyses

### SPAN and life expectancy (lifespan)

We estimated lifespan using a conventional period life table approach, modelling a hypothetical cohort exposed to age-specific mortality rates^15,50-52^. To examine the associations between SPAN behaviours and lifespan, we adjusted the mortality rates using all-cause mortality risk estimates derived from a multivariable Cox proportional hazard model (**Supplementary Methods 3**). We truncated values below the 2.5 percentile and above the 97.5 percentile for all SPAN exposures to minimise the influence of sparse or outlier data^26,39,42^. SPAN behaviours were jointly modelled as a 27-category variable representing all tertile combinations of sleep, MVPA and diet ^26^. We visualised the multivariable-adjusted associations of SPAN exposures with all-cause mortality by plotting each of the 27 categories as a forest plot with the lowest tertile of each exposure used as the reference point (**Figure 1**; event numbers shown in **Supplementary Table 2**). Models were adjusted for age, sex, ethnicity, smoking, education, Townsend deprivation index, alcohol, discretionary screen time, light intensity physical activity, medication (blood pressure, insulin, and cholesterol), previous diagnosis of major CVD, previous diagnosis of cancer, and familial history of CVD and cancer^26^. Further details on the covariates are provided in **Supplemental Table 3**. Proportional hazards assumptions were verified using Schoenfeld residuals.

**Figure 1:**
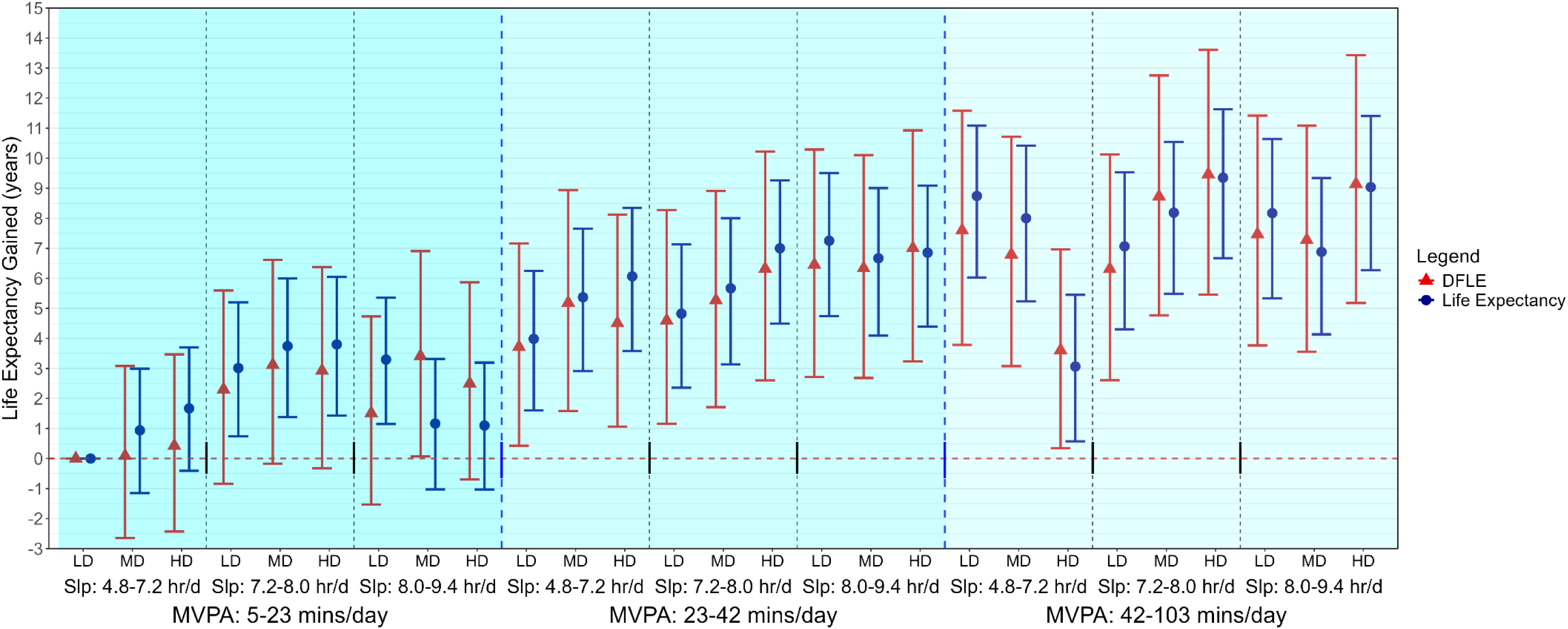
Multivariable-adjusted lifespan and healthspan associated with joint sleep, physical activity, and nutrition exposures (n = 59,078; all-cause mortality events = 2,458) Life expectancy (lifespan) was estimated using life table models, with predictions derived from hazard ratio-adjusted mortality rates for all-cause mortality associated with each category. Disease-free life expectancy (DFLE; healthspan) was calculated as the expected lifespan free from cardiovascular disease, cancer, type II diabetes, chronic obstructive pulmonary disease, or dementia. DFLE incorporated a life table approach that included age-specific incidence rates for each condition. The all-cause mortality model is adjusted for age, sex, ethnicity, smoking, education, Townsend deprivation index, alcohol, discretionary screen time (time spent watching TV or using the computer outside of work), light intensity physical activity, medication (blood pressure, insulin, and cholesterol), previous diagnosis of major CVD (defined as disease of the circulatory system, arteries, and lymph, excluding hypertension), previous diagnosis of cancer, and familial history of CVD and cancer. The specific ranges for each exposure included sleep duration as 4.8-7.2 hours/day (low), 7.2-8.0 hours/day (medium), and 8.0-9.4 hours/day (high); MVPA time: 5-23 minutes/day (low), 23-42 minutes/day (medium), and 42-103 minutes/day (high); diet quality using DQS: 32.5-50.0 (low), 50.0-57.5 (medium), and 57.5-72.5 (high). Sleep (hrs/day), physical activity (moderate to vigorous intensity – MVPA-minutes/day), and nutrition (Dietary Quality Score, DQS) were included in the model as a joint term. Dashed blue lines separate tertiles MVPA and dashed black lines separate tertiles of sleep. Sleep (Slp); Low Diet Quality (LD); Medium Diet Quality (MD); High Diet Quality (HD).

### SPAN and disease-free life expectancy (healthspan)

As per previous work^15,50^, healthspan was calculated as an extension of the original life expectancy life table approach that included age-specific baseline prevalence and incidence rates for each condition. Specifically, we calculated the lifespan free from all five major noncommunicable diseases (CVD, cancer, type 2, dementia, and COPD)^36^. We applied a Monte Carlo simulation (10,000 iterations) to account for uncertainty in disease prevalence estimates^33,35^. To reduce reverse causation, we excluded participants who died within the first year of follow-up^26,38,40^.

We assessed the potential synergistic effects of SPAN behaviours using the relative excess risk due to interaction (RERI), attributable proportion due to interaction (AP), and the synergistic effects index (S)^53^ for all-cause mortality and each chronic condition. These three indices estimate the contribution of synergistic interactions between exposures where an RERI or AP of 0 and an S value of 1 denote no interaction effect.

### Dose-response analyses and the composite SPAN score

To explore the minimum effective behaviour changes^26^ needed for meaningful improvements in lifespan and healthspan, we constructed a continuous composite SPAN score (range 0-100), with each SPAN behaviour equally weighted. Higher scores reflect closer alignment to the theoretically optimal dose, as identified from the dose-response relationship with all-cause mortality (**Supplementary Methods 4**)^54,55^. For the dose-response analyses, the 5^th^ percentile of the composite SPAN score was used as the reference group and the median was used in sensitivity analyses. We also completed additional stratified analyses for men and women^15^ where sex-specific and age-specific life tables were applied. We plotted a heatmap correlogram to illustrate lifespan gains from incremental changes in each SPAN behaviour (**Supplementary Figure 2**)^26^. No evidence of multicollinearity was found between behaviours (**Supplementary Table 4**)^56^.

### Sensitivity Analyses

We conducted several sensitivity analyses:

- Adjusted for additional sleep characteristics, including self-reported insomnia, snoring, chronotype (morning/evening person), and daytime sleepiness^57^.
- To ensure findings were robust to the choice of dietary indicator, we repeated the main analyses using the proportion of dietary ultra-processed food as a marker of diet quality (**Supplemental Table 5**)^58,59^.
- Adjusted for total energy intake and excluded participants who reported implausible sex-specific energy intake^60,61^.
- Excluded participants with poor self-rated health, smokers, participants with underweight BMI (<18.5), and those in the top 20th percentile of the frailty index
- Excluded participants with deaths in the first three years of follow-up or pre-existing chronic diseases.
- Adjusted the main models for BMI^62-64^ in the associations between SPAN exposures and life expectancy, we repeated the main analyses adjusted for BMI.

All analyses were conducted in R (version 4.4.2) using the survival, rms, and ggplot2 packages. Reporting followed the Strengthening the Reporting of Observational Studies in Epidemiology (STROBE) guidelines (**Supplemental Table 6**).

## RESULTS

### Sample

The core analytical sample comprised 59,078 participants (median age [IQR: 64.0 [7.8] years; 45.4% male). Over a median follow-up of 8.1 years, 2,458 deaths, 9,996 incident CVD, 7,681 incident cancer, 2,971 incident T2D, 1,540 incident COPD, and 508 incident dementia events occurred.

### Combined SPAN associations with lifespan and healthspan

A near-linear trend was observed across joint SPAN tertile categories, where higher levels of sleep, MVPA, and diet quality score were associated with greater gains in life expectancy and DFLE (**Figure 1**). MVPA appeared to be the primary contributor to observed gains in life expectancy and DFLE.

Compared to the theoretically worst combination of behaviours (lowest combined tertile), the minimum SPAN combination associated with improvements of lifespan and healthspan became clear once entering the moderate MVPA category (i.e., > 22 mins/day), regardless of the corresponding level of sleep and DQS. Low MVPA, moderate sleep and high diet were associated with an additional 3.80 years of lifespan (95% CI: 1.43, 6.05) and 2.93 years of healthspan (95% CI: -0.32, 6.37). In contrast, moderate MVPA, moderate sleep, and high DQS were associated with an additional 7.00 years of lifespan (95% CI: 4.49, 9.26) and 6.32 years of healthspan (95% CI: 2.60, 10.22). The highest life expectancy gained was observed with high MVPA, moderate sleep, and high DQS, which was associated with an additional 9.35 years of lifespan (95% CI: 6.67, 11.63) and 9.46 years of healthspan (95% CI: 5.45, 13.61).

### Beneficial synergistic relationship of SPAN behaviours with healthspan and lifespan

There was evidence of positive synergistic interaction between SPAN behaviours for all-cause mortality (RERI = 0.06; 95% CI: 0.005, 0.13; AP = 11.7%; 95% CI: 1-39%), and S (0.89; 95% CI: 0.84, 0.97). We observed weaker, non-significant interactions for T2D (RERI = 0.03; 95% CI: –0.01, 0.08; AP = 4.3%; 95% CI: –0.8%, 19.5%; S = 0.93; 95% CI: 0.87, 1.06) and cancer (RERI = 0.02; 95% CI: 0.00, 0.04; AP = 2.0%; 95% CI: 0%, 6.5%; S = 0.93; 95% CI: 0.88, 1.00). We found no evidence of interaction for CVD, COPD, or dementia (**Supplementary Figure 3)**.

### Dose-response associations of individual and combined SPAN behaviours with lifespan

The dose-response association was found between the composite SPAN score and lifespan (**Figure 2A)**. Compared to the 5^th^ percentile of the composite SPAN score, the median composite SPAN score (52.6) was associated with an additional 9.01 years of lifespan (95% CI: 7.31, 10.54), while a maximum score (85.3) yielded 13.14 years (95% CI: 11.41, 14.57). The estimates for the composite SPAN score and the individual behaviours stratified by sex and using the median value as the reference group are shown in **Supplementary Figure 4-5**.

**Figure 2:**
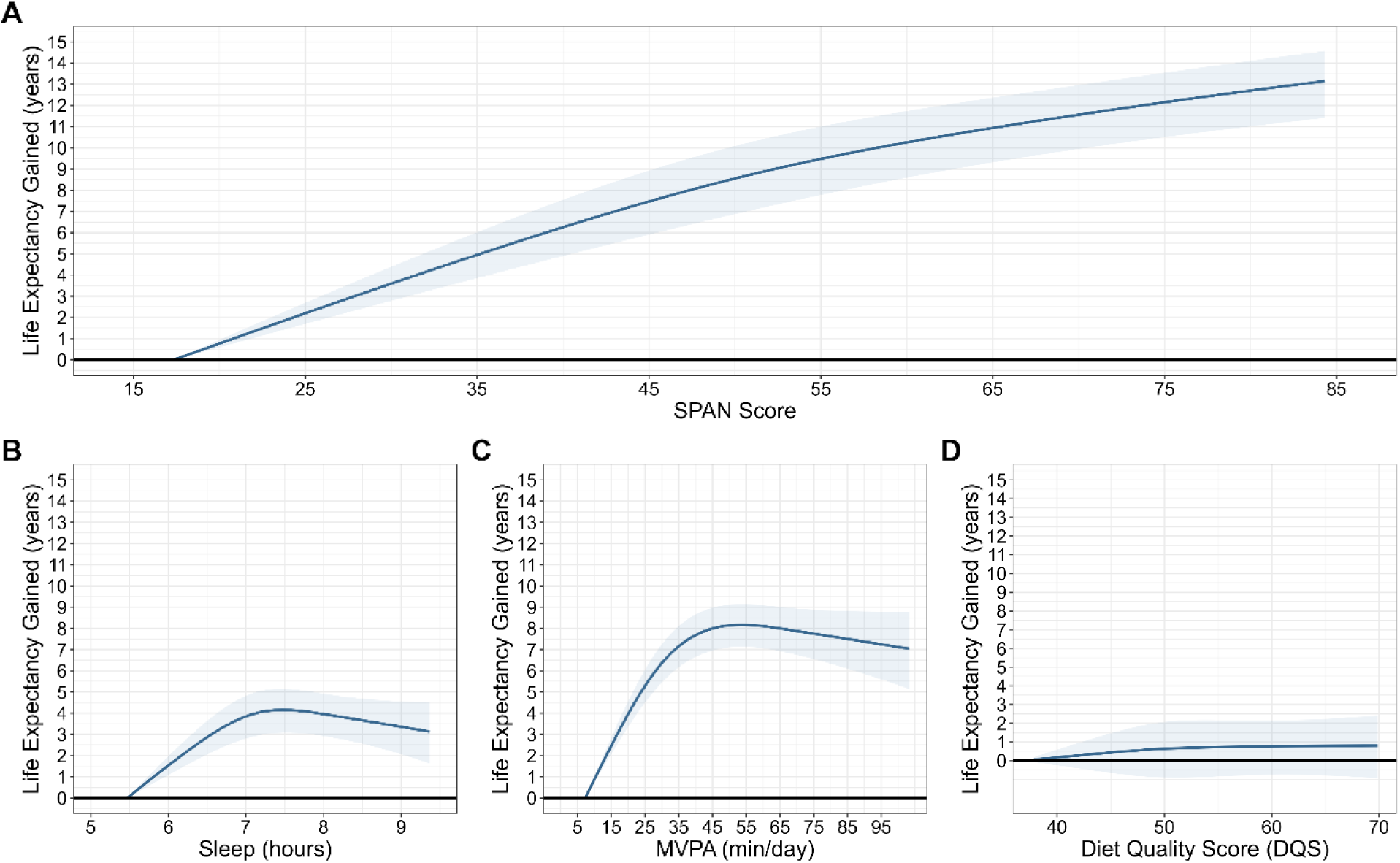
Multivariable-adjusted dose-response association between A) composite SPAN score B) sleep C) physical activity and D) nutrition with lifespan (n = 59,078; all-cause mortality events = 2,458) Life expectancy (lifespan) was estimated using stratified sex-specific life table models, with predictions based on hazard ratio-adjusted mortality rates for the association between SPAN score and all-cause mortality. The all-cause mortality model is adjusted for age, ethnicity, smoking, education, Townsend deprivation index, alcohol, discretionary screen time (time spent watching TV or using the computer outside of work), light intensity physical activity, medication (blood pressure, insulin, and cholesterol), previous diagnosis of major CVD (defined as disease of the circulatory system, arteries, and lymph, excluding hypertension), previous diagnosis of cancer, and familial history of CVD and cancer. The SPAN score is comprised of sleep (hours/day), physical activity (moderate to vigorous intensity – MVPA, minutes/day), and nutrition (Dietary Quality Score, DQS) were combined as continuous variables, each weighted equally, with scores ranging from 0 to 100. Higher scores indicated a more beneficial combined SPAN value and the referent point used was the 5^th^ percentile of the composite score. The weighting of each exposure within the SPAN score was determined based on the theoretically optimal levels identified from the dose-response relationship with all-cause mortality. In figures B-D, the individual dose-response relationship between sleep, physical activity, nutrition and life expectancy was examined using the 5^th^ percentile for each exposure as the referent point.

Sleep showed a U-shaped association with lifespan, plateauing around 7.5 hours per day (+3.39 years; 95% CI, 2.56 to 4.18: **Figure 2B**). MVPA showed a J-shaped association with lifespan, peaking at approximately 50 mins/day (+8.14 years; 95% CI, 7.11, 9.12; **Figure 2C**). The association between diet quality score and lifespan was modest and not statistically significant (**Figure 2D**).

### Minimal variations across the three SPAN behaviours and lifespan

**Table 2** presents the effects of concurrent and individual theoretical improvements in sleep, physical activity, and nutrition associated with lifespan years gained. The first rows indicate the minimal differences in the combined SPAN behaviours associated with one year of lifespan gained. For example, compared to the combined SPAN reference point (5^th^ percentile of all SPAN behaviours), an additional 5 mins/day of sleep, 1.9 mins/day MVPA, and increasing DQS by 5 points were associated with an additional 1 year of lifespan (95% CI: 0.77, 1.26). An increase of 60 mins/day of sleep, 8.8 mins/day MVPA, and 20 DQS points corresponds to a 5-increase in lifespan (95% CI: 3.98, 5.98).

**Table 1:**
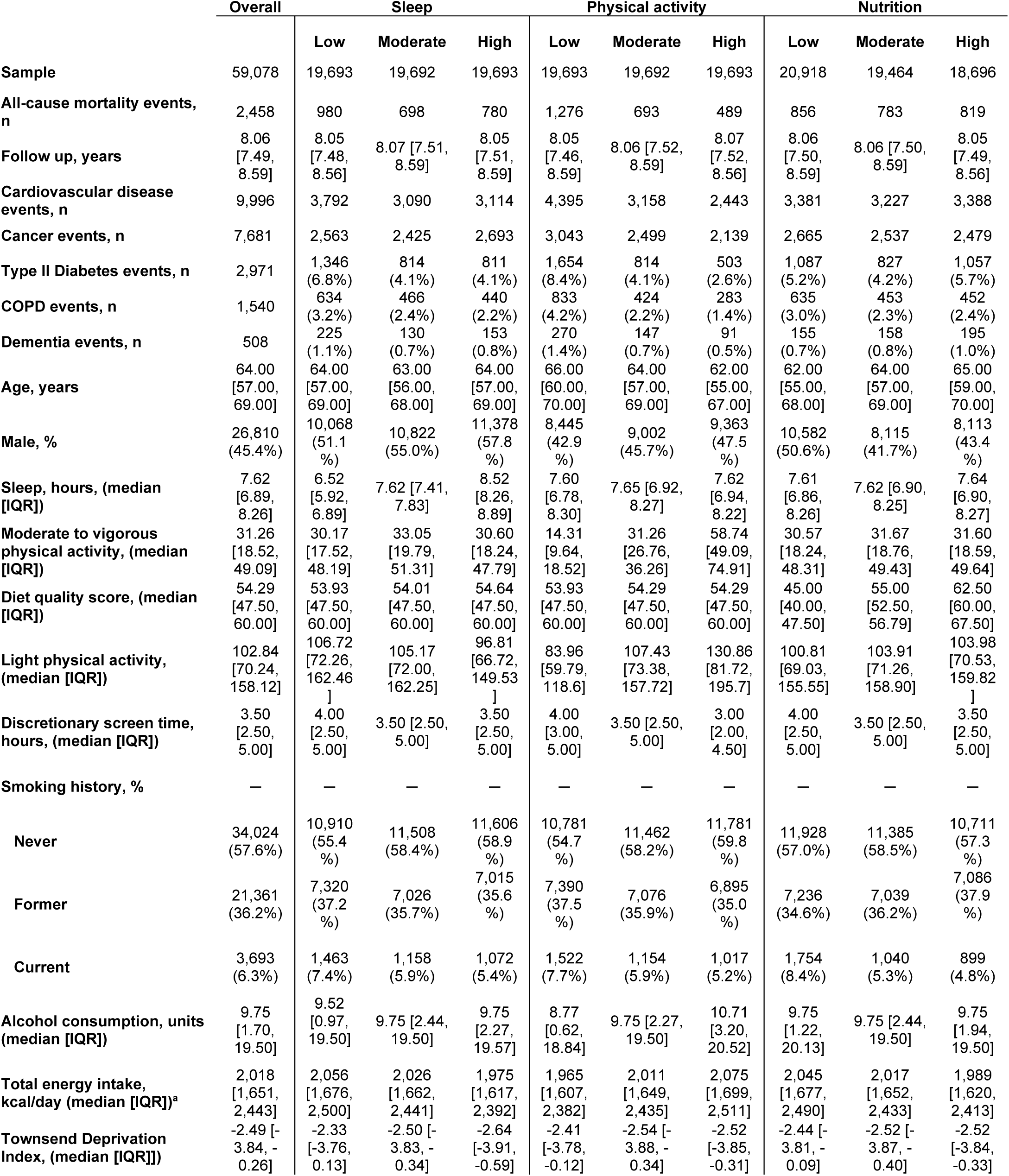
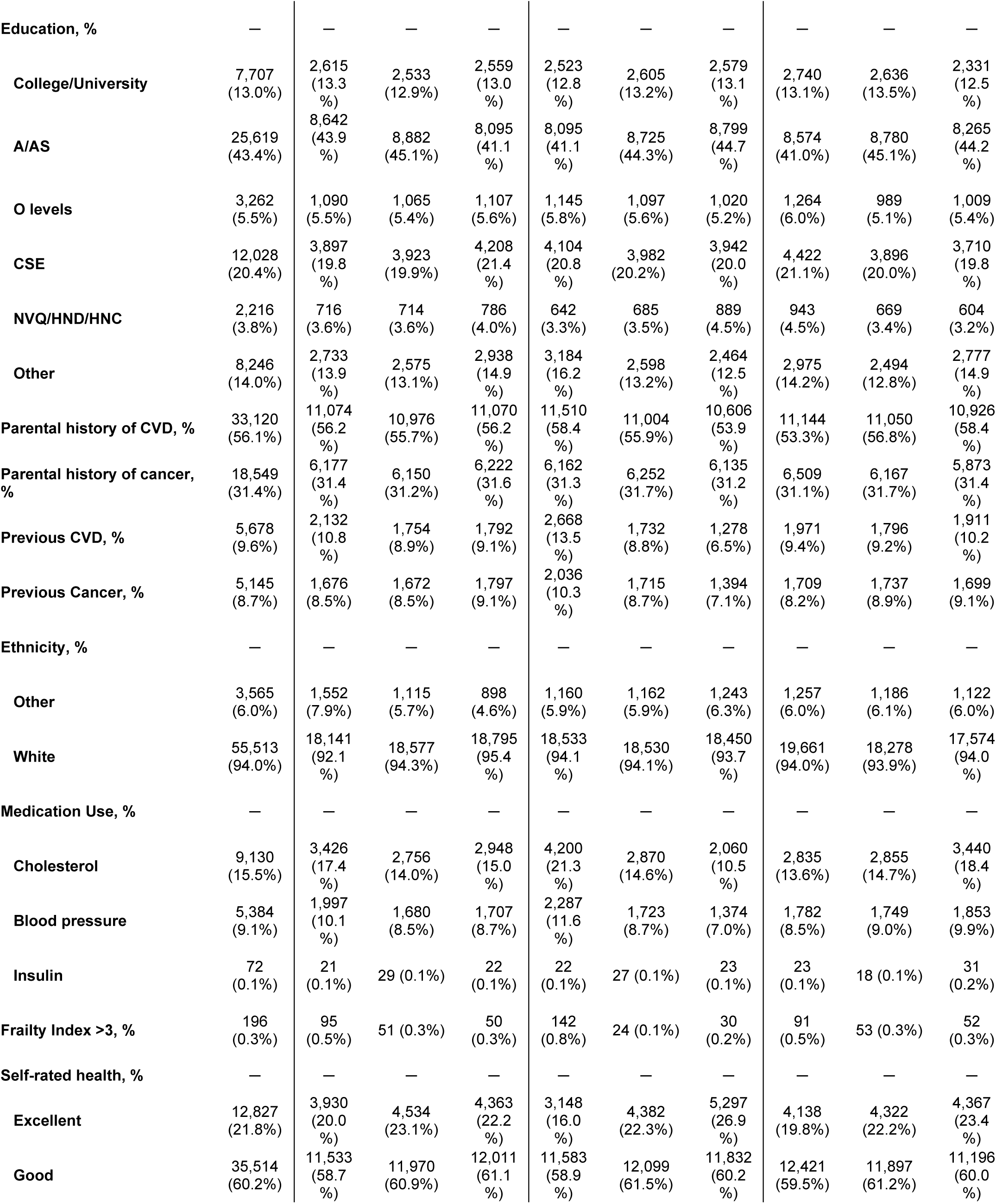
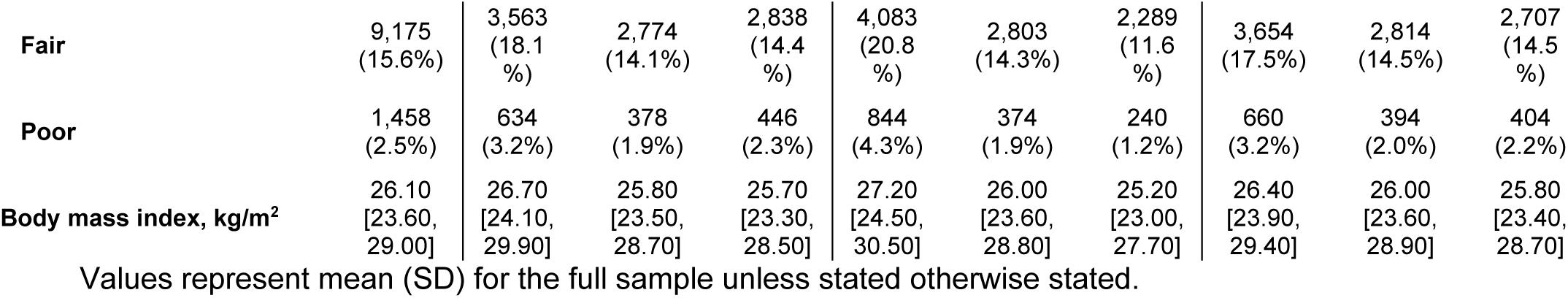
Participant characteristics.

**Table 2.** Minimum concurrent variations in sleep, physical activity, and nutrition associated with increments of lifespan gained compared to 5^th^ percentile of SPAN exposures.

| Years of Lifespan Gained<sup>§</sup> | Additional Composite SPAN Score Dose | Additional Sleep (min/day) | Additional Physical Activity (min/day) | Additional Nutrition (DQS points) | Additional Sleep* (min/day) | Additional Physical Activity* (min/day) | Additional Nutrition* (DQS points) |
| --- | --- | --- | --- | --- | --- | --- | --- |
|  |  | Combined SPAN exposure |  |  | Individual SPAN exposures |  |  |
| ≈1 Year (1.02; 95%CI: 0.77, 1.26) | 4 | 5 | 1.9 | 5 | 25 | 2.3 | 35.5 |
| ≈2 Years (2.01; 95%CI: 1.67, 2.34) | 7 | 15 | 3.8 | 5 | 49 | 4.5 | - |
| ≈3 Years (3.04; 95%CI: 2.48, 3.61) | 11 | 30 | 5.5 | 10 | 77 | 6.7 | - |
| ≈4 Years (4.02; 95%CI: 3.36, 4.67) | 14 | 45 | 7.3 | 10 | - | 9.1 | - |
| ≈5 Years (5.03; 95%CI: 3.98, 5.98) | 18 | 60 | 8.8 | 20 | - | 11.7 | - |
| ≈6 Years (6.01; 95%CI: 4.93, 7.05) | 22 | 75 | 11.0 | 20 | - | 14.8 | - |
| ≈7 Years (7.01; 95%CI: 5.60, 8.34) | 26 | 90 | 13.0 | 30 | - | 19.4 | - |
| ≈8 Years (8.01; 95%CI: 6.56, 9.36) | 30 | 120 | 15.4 | 30 | - | 47.1 | - |
| ≈9 Years (9.01; 95%CI: 7.40, 10.47) | 35 | 150 | 18.4 | 35 | - | - | - |
| ≈10 Years (10.00; 95%CI: 8.39, 11.44) | 41 | 180 | 24.9 | 35 | - | - | - |

Table displays the minimum concurrent combinations of sleep, physical, activity and nutrition associated with increments of life expectancy gained (years) compared to individual SPAN exposures. The combined columns show the SPAN combinations and corresponding years of life expectancy gained compared to the 5th percentile of sleep (5.5 hours/day), physical activity (7.3 mins/day), and nutrition (36.9 DQS) in one year increments. For comparison, the dose needed for individual SPAN exposures is shown on the right (greyed cells). Empty cells denote that the individual SPAN exposure could not achieve that gain in life expectancy in isolation. All models were adjusted for age, sex, ethnicity, smoking, education, Townsend deprivation index, alcohol, discretionary screen time (time spent watching TV or using the computer outside of work), light intensity physical activity, medication (blood pressure, insulin, and cholesterol), previous diagnosis of major CVD (defined as disease of the circulatory system, arteries, and lymph, excluding hypertension), previous diagnosis of cancer, and familial history of CVD and cancer. Moderate to vigorous physical activity (MVPA); Diet quality score (DQS).
\*Each exposure was adjusted by the median value of the other two SPAN exposures. #Here, life-expectancy gain = 0.66 years. This is the lowest adjusted HR for DQS individually. §95% CI are based on for combined SPAN model.

### Dose-response associations of individual and combined SPAN behaviours with healthspan

The composite SPAN score was linearly associated with healthspan (**Figure 3A**). The median SPAN score (52.5) was associated with an additional 4 years (3.97; 95% CI: 0.27,8.11) of healthspan. Sleep duration was associated with subtle meaningful improvement in healthspan at 8 hours/day, whereby compared to the 5^th^ percentile (5.4 hours per day) there was a gain in healthspan of 4 years (95% CI, 2.25, 4.87: **Figure 3B**), although the improvements were nullified with durations beyond 8.5 hours. Compared to the 5^th^ percentile of MVPA, there was a J-shaped association between MVPA and healthspan, with gains plateauing at approximately 75 mins/day associated with an additional 9.9 years of healthspan (95% CI, 7.62, 15.24 years gained; **Figure 3C**). There was a subtle but non-statistically significant association between DQS and healthspan (**Figure 3D**).

**Figure 3:**
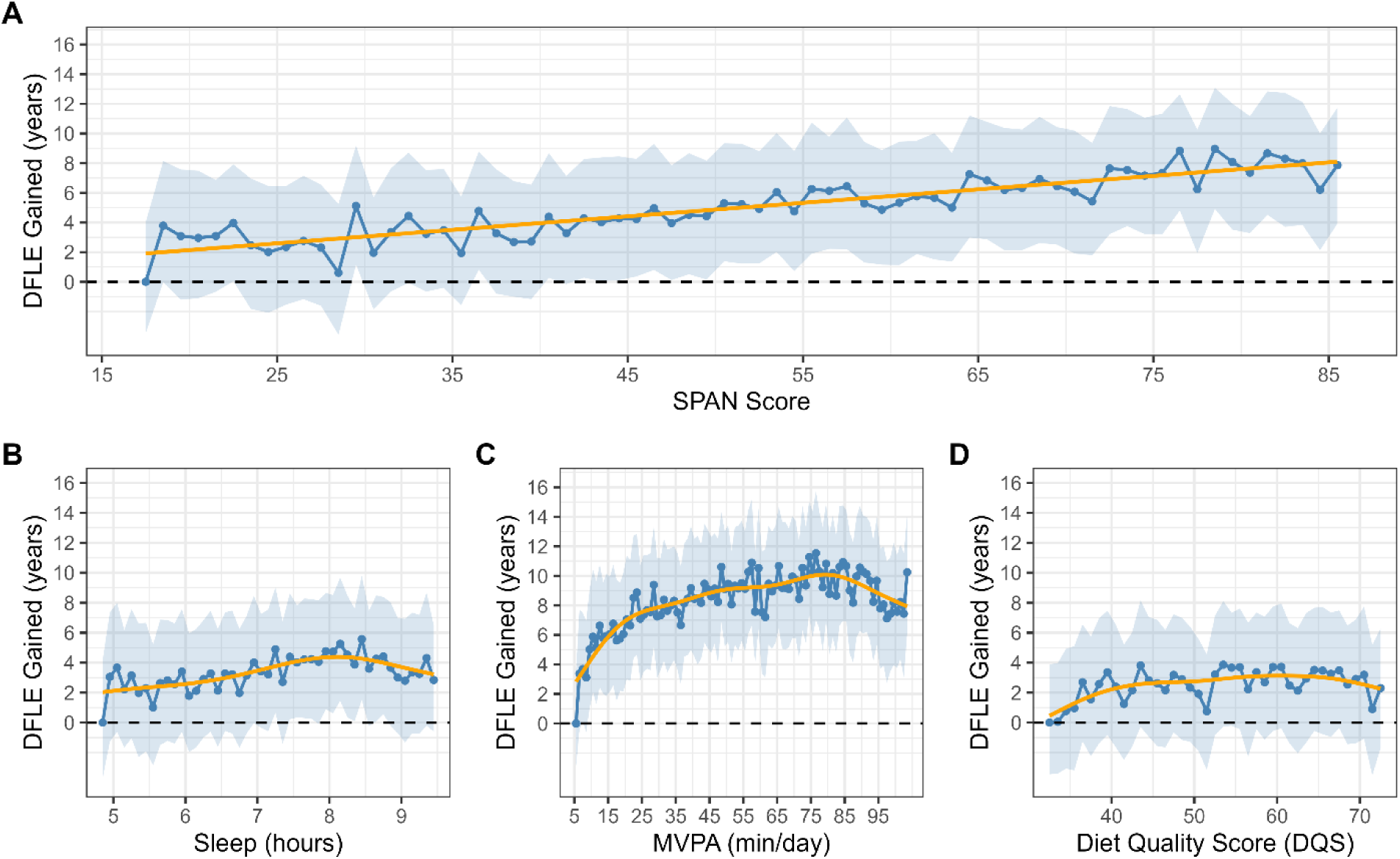
Multivariable-adjusted dose-response association between A) composite SPAN score B) sleep C) physical activity and D) nutrition with healthspan (n = 59,078; all-cause mortality events = 2,458) Disease free life expectancy (DFLE; healthspan) was estimated using stratified life table models, with predictions based on hazard ratio-adjusted mortality rates for the association between SPAN score and all-cause mortality. DFLE was calculated as an extension of the life table approach, accounting for the age-specific prevalence and incidence of the five leading contributors to disease burden, including cardiovascular disease, type II diabetes, cancer, and chronic obstructive pulmonary disease. The all-cause mortality model is adjusted for age, ethnicity, smoking, education, Townsend deprivation index, alcohol, discretionary screen time (time spent watching TV or using the computer outside of work), light intensity physical activity, medication (blood pressure, insulin, and cholesterol), previous diagnosis of major CVD (defined as disease of the circulatory system, arteries, and lymph, excluding hypertension), previous diagnosis of cancer, and familial history of CVD and cancer. The SPAN score is comprised of sleep (hours/day), physical activity (moderate to vigorous intensity – MVPA, minutes/day), and nutrition (Dietary Quality Score, DQS) were combined as continuous variables, each weighted equally, with scores ranging from 0 to 100. Higher scores indicated a more beneficial combined SPAN value, and the referent point used was the 5^th^ percentile of the composite score. The weighting of each exposure within the SPAN score was determined based on the theoretically optimal levels identified from the dose-response relationship with all-cause mortality. In figures B-D, the individual dose-response relationship between sleep, physical activity, nutrition and life expectancy was examined using the 5^th^ percentile for each exposure as the referent point.

### Minimal combined variations across the three SPAN behaviours and healthspan

**Table 3** presents the minimum theoretical combinations needed for meaningful healthspan gained. The relationship became statistically significant beyond approximately 45 points, which corresponds to nearly 4 years (3.97 years; 95% CI: 0.27, 8.11) of additional healthspan gained and achieving up to approximately 8 years (7.98 years; 95% CI: 3.47, 11.63) of additional healthspan at the maximum composite SPAN score. This minimum dose (45 points) of additional composite SPAN score is equivalent to a theoretical change of as little as 18.6 mins/day extra sleep, 3.4 mins/day MVPA, and an improvement of 21.0 DQS points (e.g. one cup of vegetables per day and two servings of fish per week). Among the individual SPAN behaviours, MVPA had the strongest relationship with healthspan (**Figure 3B**), while sleep and DQS were associated with a maximum of 5 years additional healthspan, but with uncertainty estimates that were not significant (**Figure 3B-D**).

**Table 3.** Minimum concurrent variations in sleep, physical activity, and nutrition associated with increments of healthspan gained compared to 5^th^ percentile of SPAN exposures.

| Years of Healthspan Gained* | Additional Composite SPAN Score Dose | Additional Sleep (min/day) | Additional Physical Activity (min/day) | Additional Nutrition (DQS points) | Additional Sleep<sup>†</sup> (min/day) | Additional Physical Activity<sup>†</sup> (min/day) | Additional Nutrition<sup>†</sup> (DQS points) |
| --- | --- | --- | --- | --- | --- | --- | --- |
|  |  | Combined SPAN |  |  | Individual SPAN |  |  |
| ≈4 Year (3.97; 95%CI: 0.27, 8.11) | 24 | 19 | 3.4 | 21 | 189 | 3.9 | - |
| ≈5 Year (4.97; 95%CI: 0.95, 9.10) | 35 | 28 | 5.0 | 31 | - | 6.8 | - |
| ≈6 Year (5.97; 95%CI: 1.66, 9.77) | 46 | 37 | 6.6 | 41 | - | 10.2 | - |
| ≈7 Year (6.98; 95%CI: 3.47, 11.63) | 57 | 46 | 8.3 | 50 | - | 15.1 | - |
| ≈8 Year (7.98; 95%CI: 2.39, 10.38) | 68 | 55 | 9.9 | 60 | - | 29.6 | - |

### Sensitivity Analyses

Excluding individuals with poor baseline health (n = 51,164; events = 1,887), early mortality (n = 58,610; events = 1,990), or pre-existing disease (n = 51,166; events = 1,888); adjusting for BMI (n = 58,363; events = 2,405); controlling for sleep characteristics (n = 37,475; events = 1,506) or total energy intake (n = 42,990; 1,758 events); and using an alternative diet quality metric (n = 41,936; events = 1,758) did not materially alter results (**Supplementary Figures 6–12**).

## DISCUSSION

### Main Findings

In this large prospective cohort study using a multi-behaviour analytical framework^26^, this study provides the first investigation into the minimum combined doses of device-measured sleep and physical activity, alongside a comprehensive dietary score for nutrition, associated with meaningful improvements in lifespan and healthspan. We show that while individual SPAN behaviours required substantial amounts to achieve improvements in lifespan and healthspan, when addressed in combinations, the overall dose needed for meaningful improvements was substantially lower. A minimum combined dose of an additional 5 mins/day of sleep, 1.9 mins/day of MVPA, and 5 DQS (e.g. 1.5 servings of whole grains per day) was associated with one additional year of lifespan. Similarly, an additional 18.6 mins/day of sleep, 3.4 mins/day of MVPA and an improvement of 21.0 DQS points (e.g. one cup of vegetables per day and two servings of fish per week) were associated with 4 additional years of healthspan.

Our findings build upon previous studies from the United States^28^, UK^34^ and China^50^ showing that adherence to multiple low-risk lifestyle behaviours (smoking cessation, maintaining a healthy weight, improved diet quality, and reduced alcohol intake) can increase lifespan by 8 to 14 years and extended years healthspan for nearly a decade in both men and women^34^. However, prior studies^27,29-33,35^ relied almost exclusively on self-reported questionnaire data, which were often assessed in isolation, and have not explored synergistic interactions between behaviors^65,66^. Here, using device-based measurements for sleep and physical activity in combination with a comprehensive diet quality index, we show the most optimal behavioural combinations linked to nearly a decade of additional lifespan and healthspan.

Our findings for physical activity are consistent with previous UK Biobank data, which have shown a gain of approximately 2 years of additional years of lifespan in those self-reporting high levels of total physical activity (≥3000 MET-min per week^67^, equivalent to approximately 12.5 hours of brisk walking or 5 hrs of running). In contrast, meeting the physical activity guidelines (≈ 22 mins/day of MVPA) through device-based measures was associated with a 5-year gain in life expectancy, consistent with our findings (≈ 19 mins/day of MVPA in isolation)^67^. Sleep has also shown a strong association with lifespan^57,68,69^ and healthspan^70^. Our results reinforce growing evidence of its foundational role in metabolic health^4,5,24^, showing a maximum gain of ≈4 years additional lifespan and health span when sleeping ≈8 hrs/day compared to the 5^th^ percentile (5 hrs/day). While dietary associations remained subtle and non-significant in isolation, the combination of diet with sleep and physical activity suggests it plays an important synergistic role, as supported by the RERI. Longer follow-up with repeated measures to more fully characterise the contribution of diet to lifespan and healthspan.

A central finding of our study is the demonstration of synergistic effects of SPAN behaviours with healthspan and lifespan. This is biologically plausible and grounded in the unique interconnected physiological pathways between these behaviours^26,71-73^, including circadian alignment, energy regulation, and metabolic adaptations^4,5,24^. When considered individually, substantially higher levels of each behaviour were required to achieve a similar extension in lifespan, or the improvements were not possible at all. For example, to theoretically gain one additional year of lifespan would require 25 minutes of additional sleep per day, with a maximum lifespan gain of three years. In combination, however, relatively modest changes, such as 5 additional mins/day of sleep, 1.9 mins/day of MVPA, and half a serving of vegetables per day, were associated with a meaningful extension of lifespan by one year. For healthspan, this was particularly evident, whereby physical activity required exponentially higher doses to achieve the same healthspan benefit as the combined SPAN behaviours. For example, 29.6 mins/day MVPA was necessary to achieve 8 additional years free of chronic disease vs. 9.9 mins/day MVPA per day in combination with sleep (55 additional mins/day) and dietary improvements (60 additional DQS points). These findings suggest that leveraging the synergy of combined SPAN behaviours may enable new opportunities for feasible and sustainable approaches for improving lifespan and healthspan in adults who may lack the time, motivation, or financial resources to make substantial lifestyle changes.

### Strengths and limitations

To our knowledge, this is the first study to apply a high-resolution, device-based, multi-behaviour analytical approach^26^ to estimate the minimum combined improvements in SPAN behaviours associated with healthspan and lifespan. Previous work exploring the role of lifestyle behaviours with lifespan^27-31,33,34^ has done so through broad self-reported categorical terms, limiting the translation into feasible and practical messaging for behaviour change. By modelling behaviour changes in interpretable units (e.g., mins/day, servings/week), we provide directly actionable insights for public health messaging and behavioural interventions. The use of accelerometry-derived data for sleep and physical activity is a major strength, offering improved measurement precision over self-report.

Furthermore, by including participants from the UK Biobank’s wearables sub-study, we were able to incorporate device-based estimates for both sleep and physical activity, offering a more accurate and granular assessment of these behaviours. However, the measurement source of sleep and physical activity presents a limitation when making comparisons with nutrition, which was self-reported in this study. This methodological difference may partially explain why the findings differ from existing literature on the association between diet with lifespan and healthspan^28,33,35^, as we observed a subtle non-significant relationship. This discrepancy may reflect differences in measurement quality, where diet may have been more significantly impacted by regression dilution bias^74^, attenuating its true association with mortality and morbidity. In the UK Biobank^37,48^, there is also a temporal lag between the data collection of the behaviours, as FFQ dietary data were collected a median of 5.5 years before the wearable device measures of sleep and physical activity. The possibility of reverse causation or residual confounding cannot be eliminated, although we conducted a range of sensitivity analyses, including the exclusion of participants with a mortality event within the first three years of follow-up, self-rated poor health, high frailty index, underweight BMI, or a chronic condition at baseline. Future longitudinal and interventional research is needed to confirm these findings, particularly to understand the sustainability of small SPAN behaviour changes, the time required to achieve meaningful health effects, and the impact of SPAN behaviour change duration. While other unhealthy behaviours, such as smoking and alcohol, were accounted for in this study, further research is required to examine their interactions with SPAN behaviours, their interrelationship with the chronic conditions examined, and their impact on lifespan and healthspan.

## CONCLUSIONS

This study demonstrates that small, combined improvements in sleep, physical activity, and nutrition are associated with meaningful increases in both lifespan and healthspan. Gains of one additional year of lifespan were achievable through relatively modest combined changes, such as an additional 5 mins/day of sleep per day, just under 2 mins/day MVPA, and half a serving of vegetables per day. For healthspan, a theoretical minimum change of 18.6 mins/day of extra sleep, 3.4 mins/day of MVPA and extra 21.0 DQS points (e.g. an additional 1 cup of vegetables per day and two servings per week of fish) was associated with a clinically meaningful gain of four additional years free of major chronic conditions. These findings suggest that small, achievable, combined changes in SPAN behaviours, may offer a powerful and feasible public health opportunity for improving healthspan and lifespan.

## Data Availability

The data that support the findings of this study are available from the UK Biobank but restrictions apply to the availability of these data, which were used under license for the current study, and so are not publicly available. Data are however available from the authors upon reasonable request and with permission of the UK Biobank.

## Acknowledgements

This research has been conducted using the UK Biobank resource under application number 25813. The authors would like to thank all the participants and professionals contributing to the UK Biobank. All information and materials in the manuscript are original and have not been submitted for publication elsewhere.

## Data availability Statement

The data that support the findings of this study are available from the UK Biobank, but restrictions apply to the availability of these data, which were used under license for the current study and so are not publicly available. Data are, however, available from the authors upon reasonable request and with the permission of the UK Biobank.

