## Supplementary Material for "Minimum concurrent sleep, physical activity, and nutrition variations associated with lifeSPAN and healthSPAN improvements: a population cohort study"


| Page | Item |
|---|---|
| **3** | **Supplemental Figure 1:** Flow diagram of participants in the study |
| **4** | **Supplementary Figure 2:** Years of lifespan gained with concurrent variations in sleep, physical activity, and nutrition (n = 59,078; all-cause mortality events = 2,458) |
| **5** | **Supplementary Figure 3:** Synergistic relationship between sleep, physical activity, and nutrition in relation to mortality and morbidity |
| **6** | **Supplementary Figure 4:** Multivariable-adjusted dose-response association between A) SPAN score B) sleep C) physical activity and D) nutrition with lifespan in males (n = 26,810; all-cause mortality events = 1,520) and females (n = 32,268; all-cause mortality events = 938) |
| **7** | **Supplementary Figure 5:** Multivariable-adjusted dose-response association between A) SPAN score B) sleep C) physical activity and D) nutrition with lifespan in males (n = 26,810; all-cause mortality events = 1,520) and females (n = 32,268; all-cause mortality events = 938) using alternative referent point |
| **8** | **Supplemental Figure 6:** Multivariable-adjusted associations of combined sleep, physical activity, and nutrition with lifespan and healthspan following exclusion of individuals with poor health (n = 51,164; events = 1,887) |
| **9** | **Supplemental Figure 7:** Multivariable-adjusted associations of combined sleep, physical activity, and nutrition with lifespan and healthspan excluding individuals with baseline chronic conditions (n = 51,166; events = 1,888) |
| **10** | **Supplemental Figure 8:** Multivariable-adjusted associations of combined sleep, physical activity, and nutrition with lifespan and healthspan excluding individuals with a mortality event in the first three years of follow-up (n = 58,610; events = 1,990) |
| **11** | **Supplemental Figure 9:** Multivariable-adjusted associations of combined sleep, physical activity, and nutrition with lifespan and healthspan adjustment for BMI (n = 58,363; events = 2,405) |
| **12** | **Supplemental Figure 10:** Multivariable-adjusted associations of combined sleep, physical activity, and nutrition with lifespan and healthspan adjustment for sleep characteristics (n = 37,475; events = 1,506) |
| **13** | **Supplemental Figure 11:** Multivariable-adjusted associations of combined sleep, physical activity, and nutrition with lifespan and healthspan using the proportion of ultra-processed food (n = 41,936; events = 1,758) |

| | |
|---|---|
| 14 | **Supplemental Figure 12:** Multivariable-adjusted associations of combined sleep, physical activity, and nutrition with lifespan and healthspan adjusted for total energy intake (n = 42,990; 1,758 events) |
| 15 | **Supplementary Methods 1:** Additional study design details |
| 18 | **Supplementary Methods 2:** Wearable behaviour classification methods |
| 21 | **Supplementary Methods 3:** Calculation of lifespan and healthspan |
| 24 | **Supplementary Methods 4:** Calculation of the composite SPAN score |
| 25 | **Supplemental Table 1:** Diet quality score index for food-frequency questionnaire dietary data |
| 26 | **Supplemental Table 2:** Mortality and disease events across the mutually exclusive sleep, physical activity, and nutrition combinations |
| 27 | **Supplemental Table 3:** Covariate definitions |
| 29 | **Supplementary Table 4:** Model variance inflation factors for combined SPAN behaviours |
| 30 | **Supplementary Table 5:** NOVA classification of food groups for 24-hour dietary recall data |
| 31 | **Supplemental Table 6:** STROBE statement |

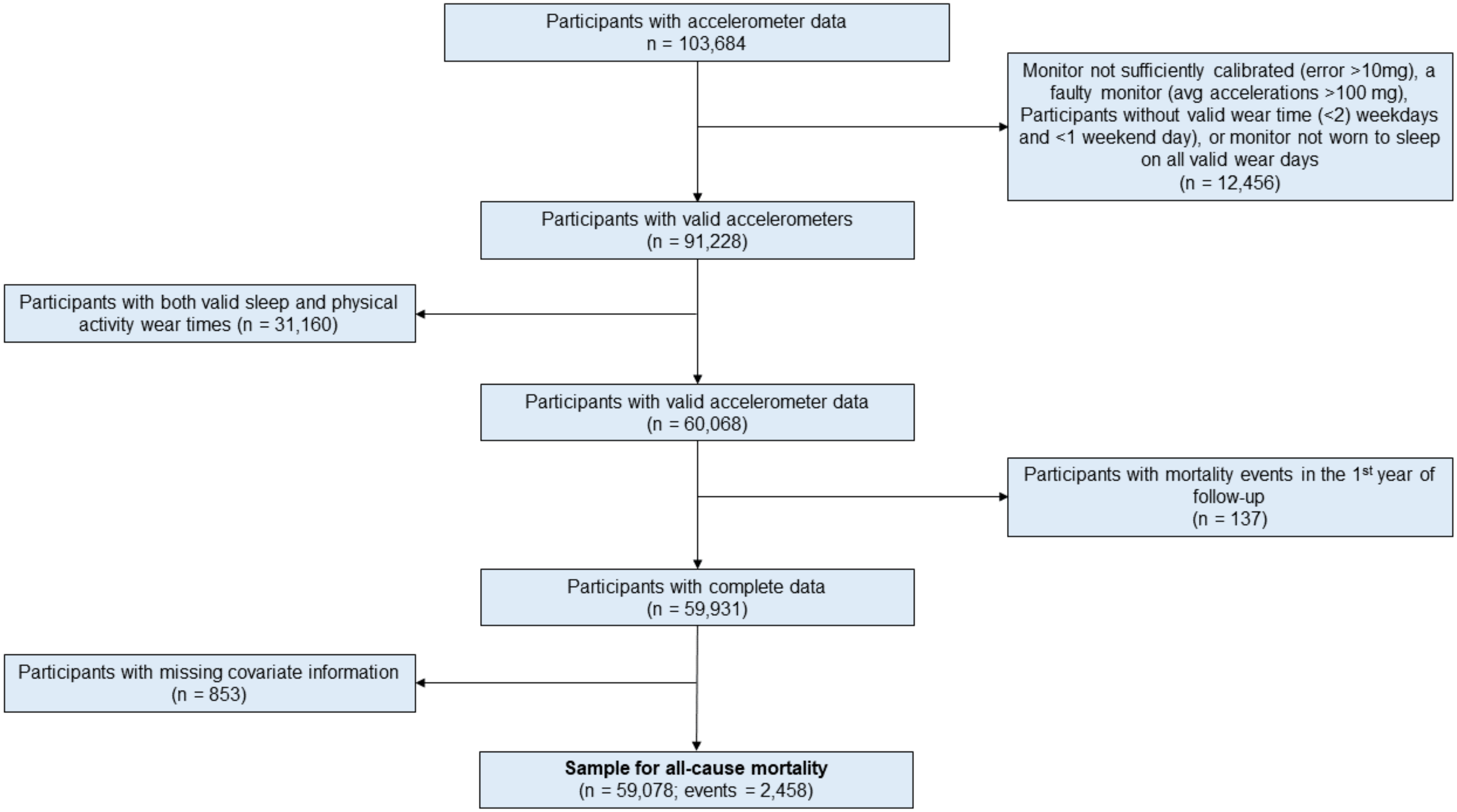


**Supplementary Figure 1.** Participant flow chart

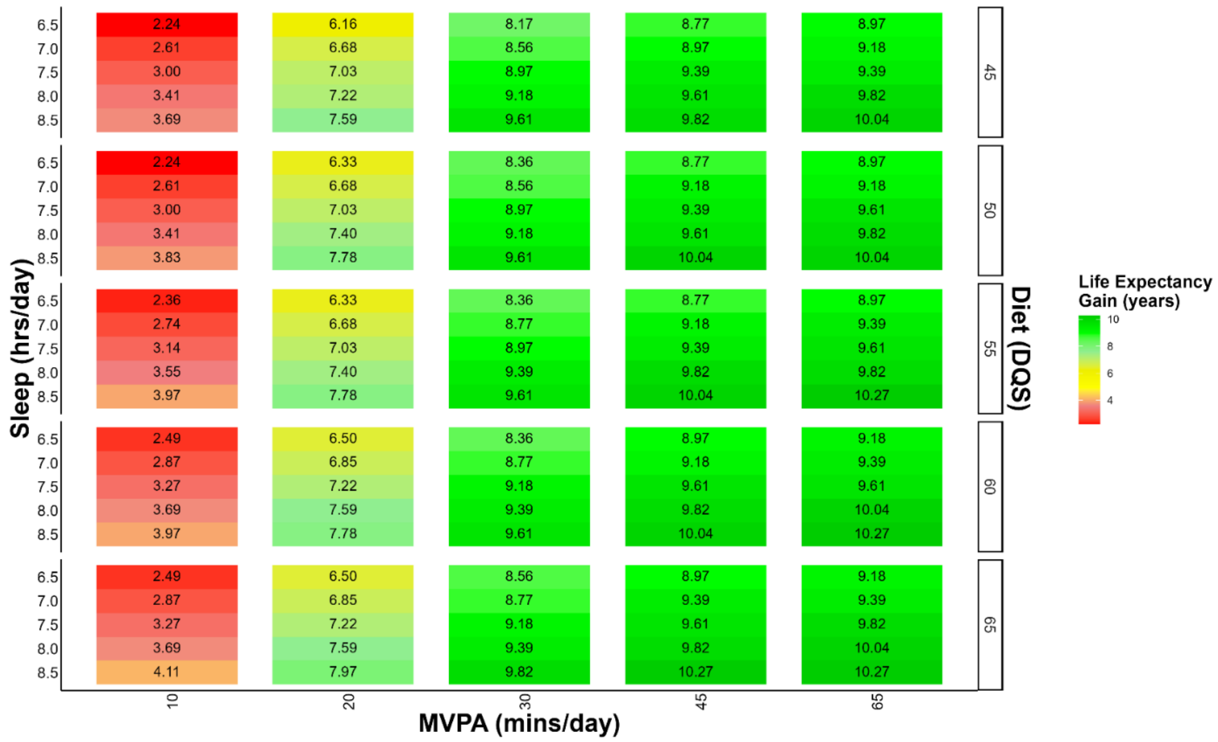


**Supplementary Figure 2.** Years of lifespan gained with concurrent variations in sleep, physical activity, and nutrition (n = 59,078; all-cause mortality events = 2,458)

**Legend:** The correlogram displays changes in sleep (hrs/day), physical activity (moderate to vigorous intensity (MVPA) minutes/day), and nutrition (Dietary Quality Score (DQS)) and corresponding years of life expectancy (lifespan) gained with the reference being the 5th percentile of sleep (5.5 hours/day), physical activity (7.3 minutes/day), and nutrition (36.9 DQS). Sleep, physical activity, and nutrition are included as independent terms in the model to allow for more granular predictions. Life expectancy was estimated using life table models, with predictions based on hazard ratio-adjusted mortality rates for all-cause mortality. The all-cause mortality is adjusted for age, sex, ethnicity, smoking, education, Townsend deprivation index, alcohol, discretionary screen time (time spent watching TV or using the computer outside of work), light intensity physical activity, medication (blood pressure, insulin, and cholesterol), previous diagnosis of major CVD (defined as a disease of the circulatory system, arteries, and lymph, excluding hypertension), previous diagnosis of cancer, and familial history of CVD and cancer.

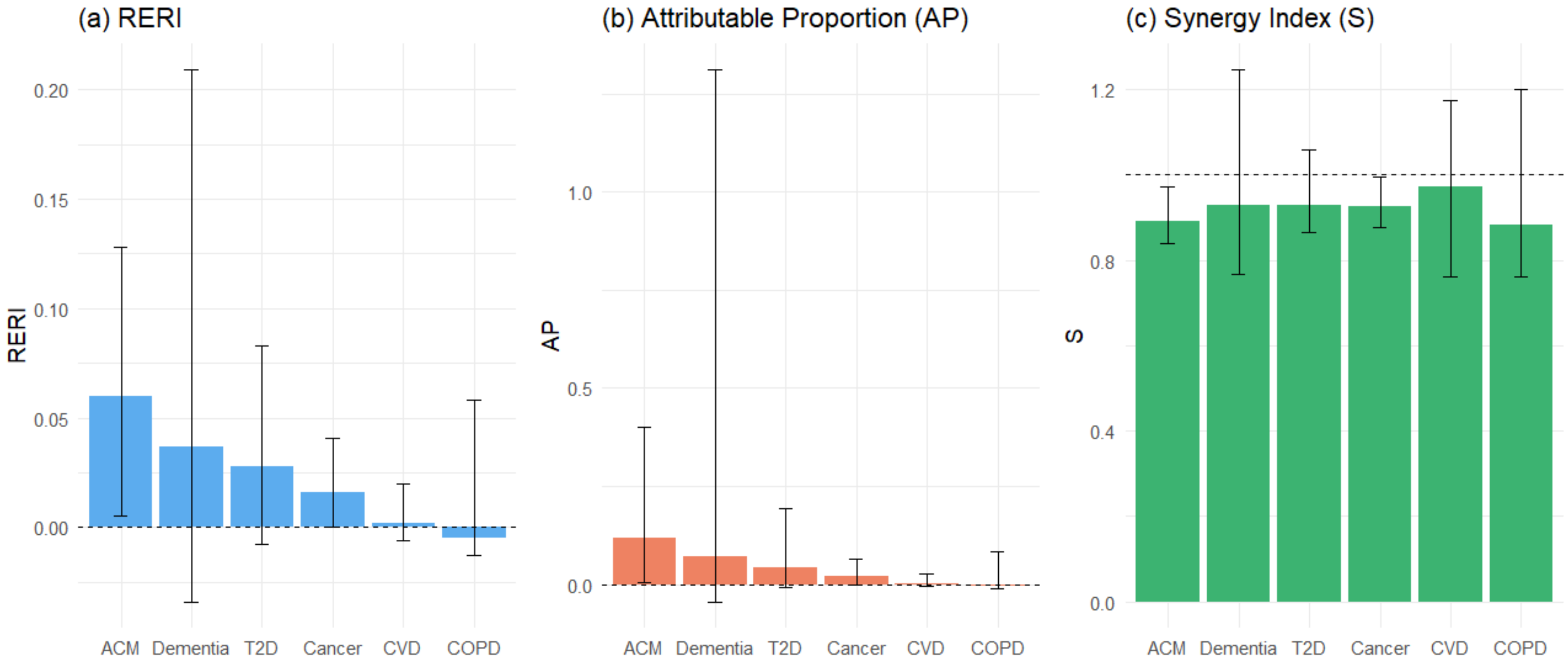


**Supplementary Figure 3**. Synergistic relationship between sleep, physical activity, and nutrition in relation to mortality and morbidity

**Legend**: The figure above shows the individual and interactive model terms of sleep, physical activity, and nutrition for all-cause mortality (ACM); type II diabetes (T2D); cardiovascular disease (CVD), and chronic obstructive pulmonary disease (COPD). To test for interactive and synergistic effects, we calculated the relative excess risk due to interaction (RERI), attributable proportion due to interaction (AP), and the synergistic effects index (S)[1]. These tests provide insight into the contribution of synergistic interactions between exposures where an RERI or AP of 0 and an S value of 1 denote no interaction effect.

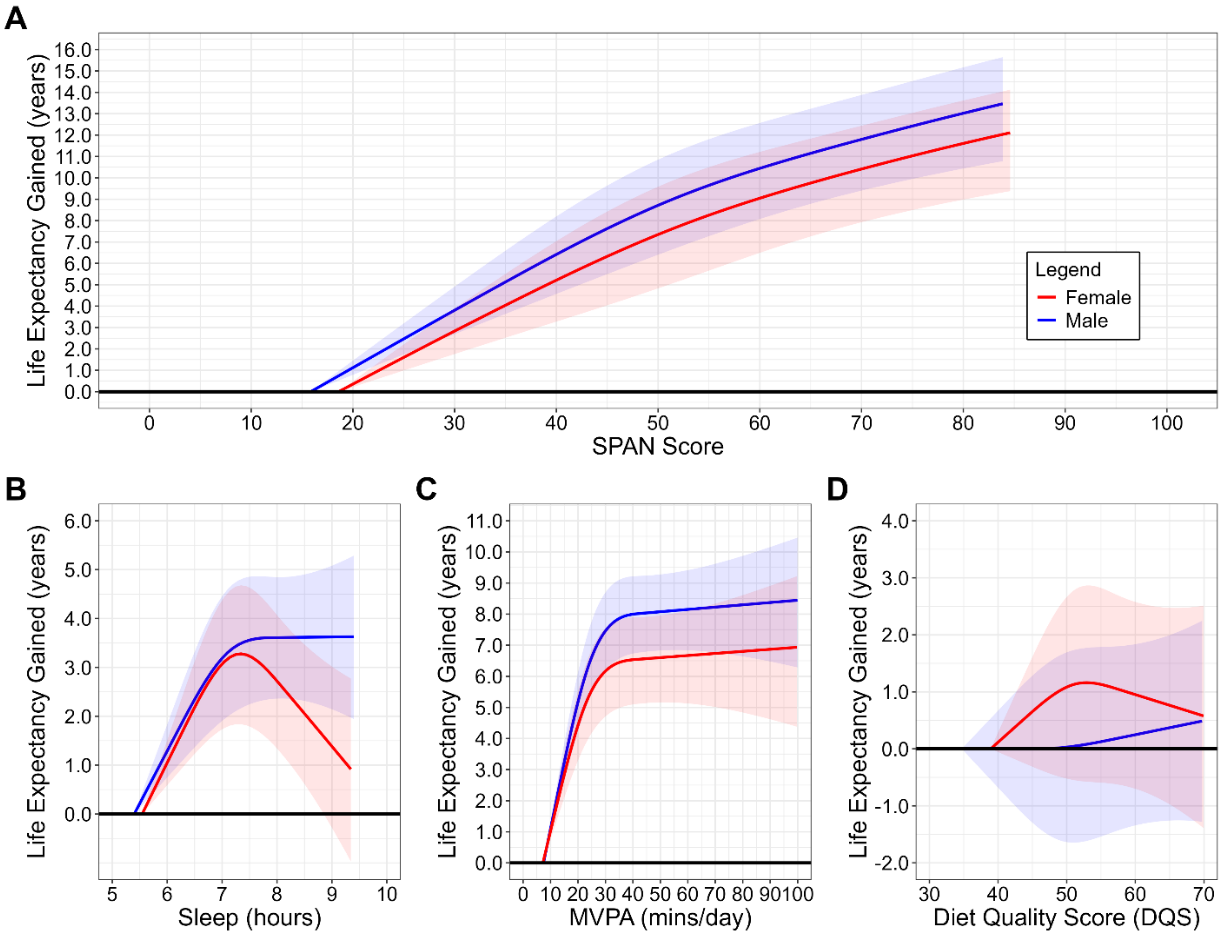


**Supplementary Figure 4.** Multivariable-adjusted dose-response association between A) SPAN score B) sleep C) physical activity and D) nutrition lifespan in males (n = 26,810; all-cause mortality events = 1,520) and females (n = 32,268; all-cause mortality events = 938)

**Legend:** Life expectancy was estimated using stratified sex-specific life table models, with predictions based on hazard ratio-adjusted mortality rates for the association between SPAN score and all-cause mortality. The all-cause mortality model is adjusted for age, ethnicity, smoking, education, Townsend deprivation index, alcohol, discretionary screen time (time spent watching TV or using the computer outside of work), light intensity physical activity, medication (blood pressure, insulin, and cholesterol), previous diagnosis of major CVD (defined as disease of the circulatory system, arteries, and lymph, excluding hypertension), previous diagnosis of cancer, and familial history of CVD and cancer. The SPAN score is comprised of sleep (hours/day), physical activity (moderate to vigorous intensity – MVPA, minutes/day), and nutrition (Dietary Quality Score, DQS) were combined as continuous variables, each weighted equally, with scores ranging from 0 to 100. Higher scores indicated a more beneficial combined SPAN value and the referent point used was the 5$^{th}$ percentile for each sex. The weighting of each exposure within the SPAN score was determined based on the theoretically optimal levels identified from the dose-response relationship with all-cause mortality. In figures B-D, the individual dose-response relationship between sleep, physical activity, nutrition and life expectancy was examined using the sex-specific 5$^{th}$ percentile for each exposure as the referent point.

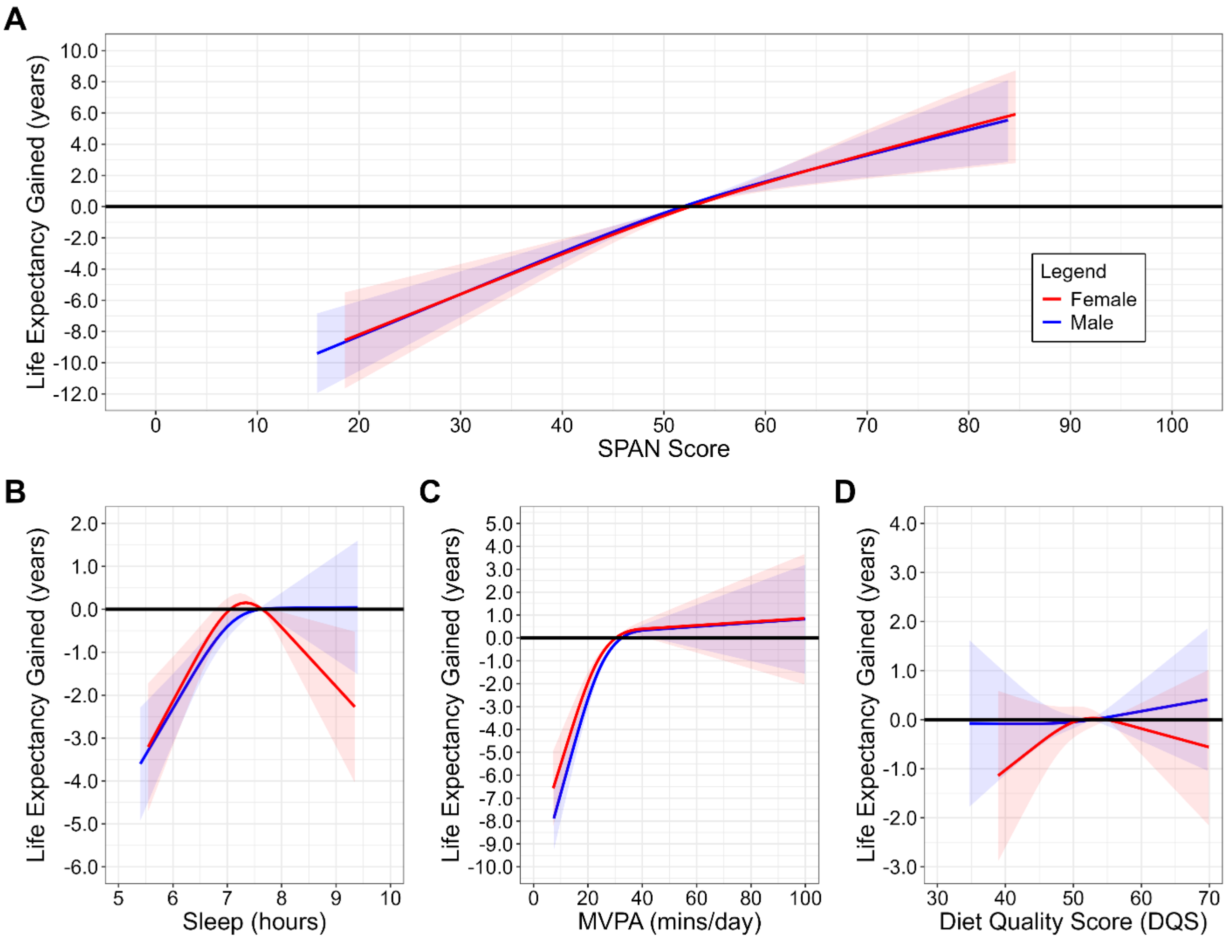


**Supplementary Figure 5.** Multivariable-adjusted dose-response association between B) sleep C) physical activity and D) nutrition with lifespan in males (n = 26,810; all-cause mortality events = 1,520) and females (n = 32,268; all-cause mortality events = 938) using alternative referent point

**Legend:** Life expectancy (lifespan) was estimated using stratified sex-specific life table models, with predictions based on hazard ratio-adjusted mortality rates for the association between SPAN score and all-cause mortality. The all-cause mortality model is adjusted for age, ethnicity, smoking, education, Townsend deprivation index, alcohol, discretionary screen time (time spent watching TV or using the computer outside of work), light intensity physical activity, medication (blood pressure, insulin, and cholesterol), previous diagnosis of major CVD (defined as disease of the circulatory system, arteries, and lymph, excluding hypertension), previous diagnosis of cancer, and familial history of CVD and cancer. The SPAN score is comprised of sleep (hours/day), physical activity (moderate to vigorous intensity – MVPA, minutes/day), and nutrition (Dietary Quality Score, DQS) were combined as continuous variables, each weighted equally, with scores ranging from 0 to 100. Higher scores indicated a more beneficial combined SPAN value and the referent point used was the median value for each sex. The weighting of each exposure within the SPAN score was determined based on the theoretically optimal levels identified from the dose-response relationship with all-cause mortality. In figures B-D, the individual dose-response relationship between sleep, physical activity, nutrition and life expectancy was examined using the sex-specific median value for each exposure as the referent point.

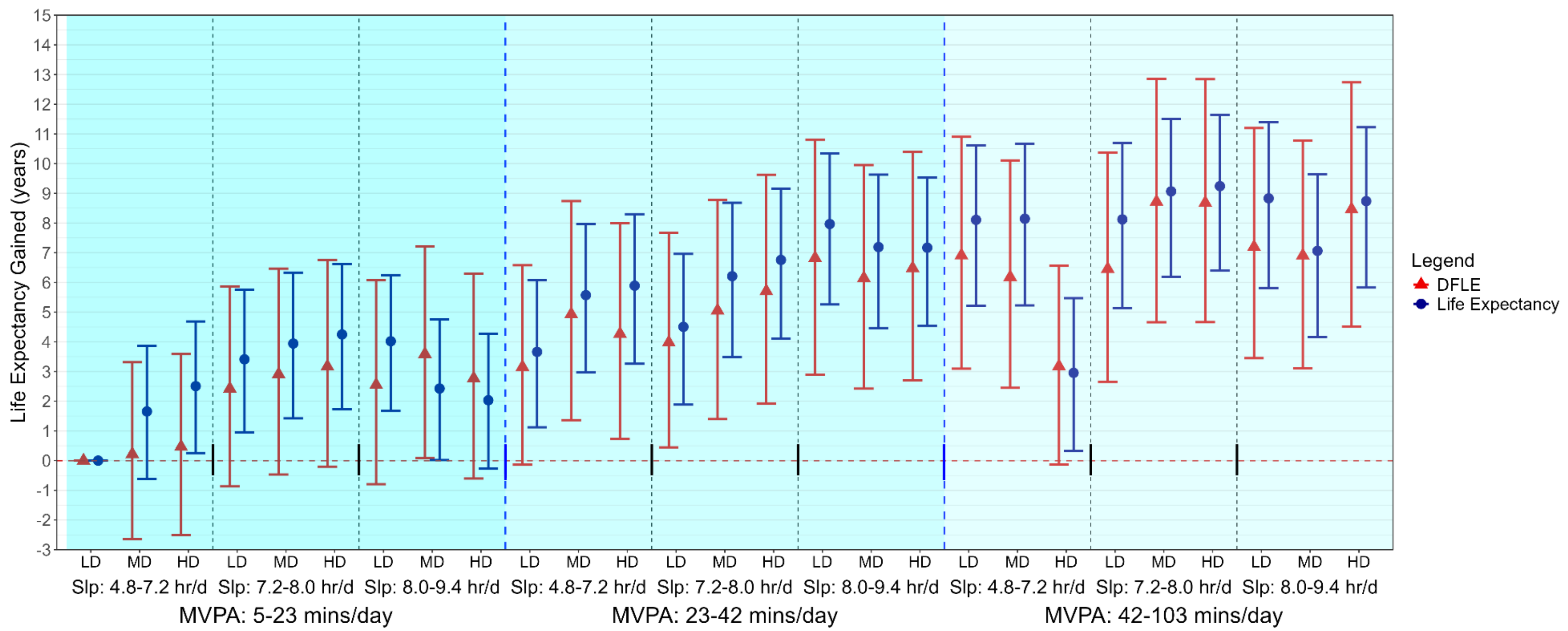


**Supplemental Figure 6:** Multivariable-adjusted associations of combined sleep, physical activity, and nutrition with lifespan and healthspan expectancy following exclusion of individuals with poor health (n = 51,164; events = 1,887)

**Legend**: Forest plot shows the SPAN associations with life expectancy (lifespan) and disease free life expectancy (DFLE; healthspan) after removing those with poor health status including low BMI (<18.5), current smokers, self-reported poor health, and those with a frailty index score of >3. Life expectancy was estimated using life table models, with predictions based on hazard ratio-adjusted mortality rates for all-cause mortality associated with each category. Disease-free life expectancy was calculated as the expected lifespan free from cardiovascular disease, cancer, type II diabetes, chronic obstructive pulmonary disease (COPD), or dementia. DFLE incorporated a life table approach that included age-specific incidence rates for each condition. The all-cause mortality is adjusted for age, sex, ethnicity, smoking, education, Townsend deprivation index, alcohol, discretionary screen time (time spent watching TV or using the computer outside of work), light intensity physical activity, medication (blood pressure, insulin, and cholesterol), previous diagnosis of major CVD (defined as disease of the circulatory system, arteries, and lymph, excluding hypertension), previous diagnosis of cancer, and familial history of CVD and cancer. Sleep (hrs/day), physical activity (moderate to vigorous intensity – MVPA- minutes/day), and nutrition (Dietary Quality Score, DQS) were included in the model as a joint term. Dashed blue lines separate tertiles MVPA and dashed black lines separate tertiles of sleep. Sleep (Slp); Low Diet Quality (LD); Medium Diet Quality (MD); High Diet Quality (HD).

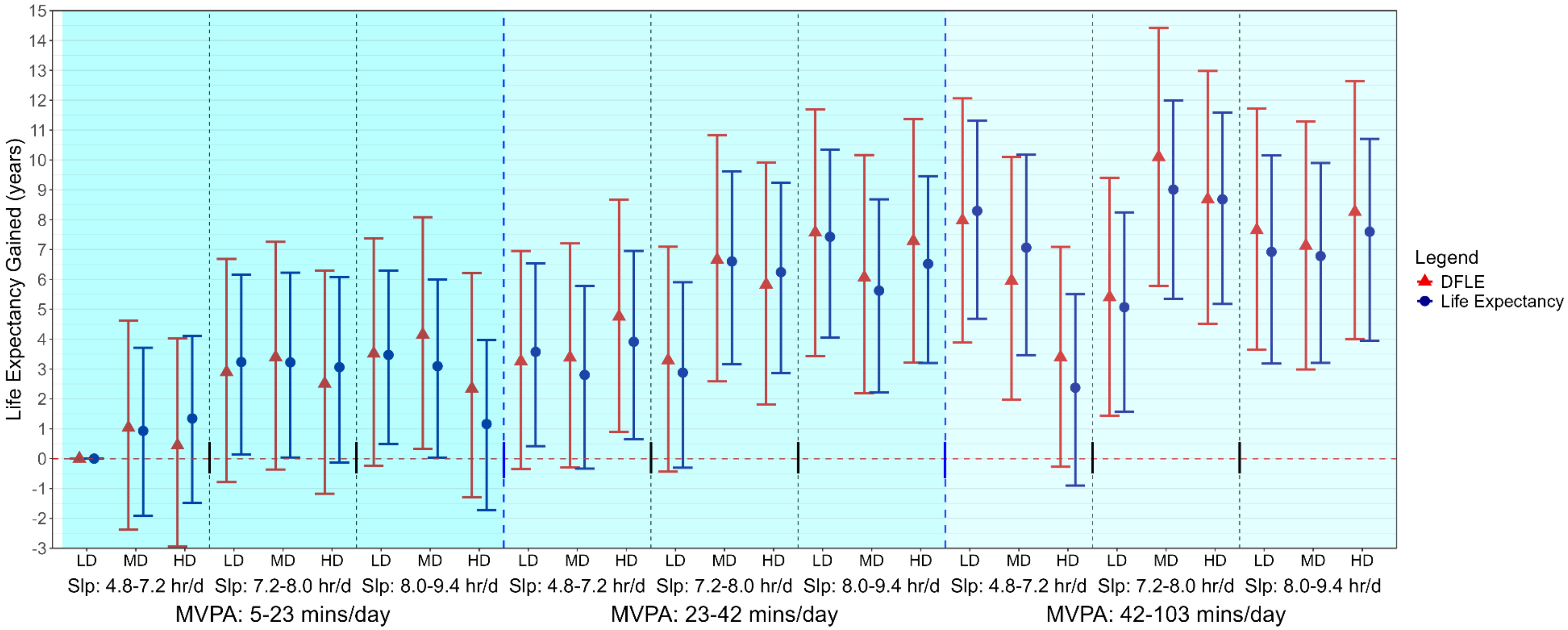


**Supplementary Figure 7:** Multivariable-adjusted associations of combined sleep, physical activity, and nutrition with lifespan and healthspan excluding individuals with baseline chronic conditions (n = 51,166; events = 1,888)

**Legend**: Forest plot shows the SPAN associations with life expectancy (lifespan) and disease free life expectancy (DFLE; healthspan) after removing those with baseline chronic conditions including cardiovascular disease (CVD), cancer, chronic obstructive pulmonary disease (COPD), dementia, or type II diabetes. Model is adjusted for age, sex, ethnicity, smoking, education, Townsend deprivation index, alcohol, discretionary screen time (time spent watching TV or using the computer outside of work), light intensity physical activity, medication (blood pressure, insulin, and cholesterol), and familial history of CVD and cancer. Sleep (hours/day), physical activity (moderate to vigorous intensity (MVPA) minutes/day), and nutrition (Dietary Quality Score (DQS)) were included in the model as a joint term. The specific ranges for each exposure included sleep duration as 5.0-7.2 hours/day (low), 7.2-8.0 hours/day (medium), and 8.0-9.4 hours/day (high); MVPA measurements as 6-23 minutes/day (low), 23-42 minutes/day (medium), and 43-104 minutes/day (high); and diet quality using the DQS as 34.0-50.0 (low), 50.0-57.5 (medium), and 57.5-72.5 (high). The lowest tertiles for all three exposures (sleep, MVPA and DQS) was the referent group. Dashed blue lines separate tertiles MVPA and dashed black lines separate tertiles of sleep. Sleep (Slp); Low Diet Quality (LD); Medium Diet Quality (MD); High Diet Quality (HD).

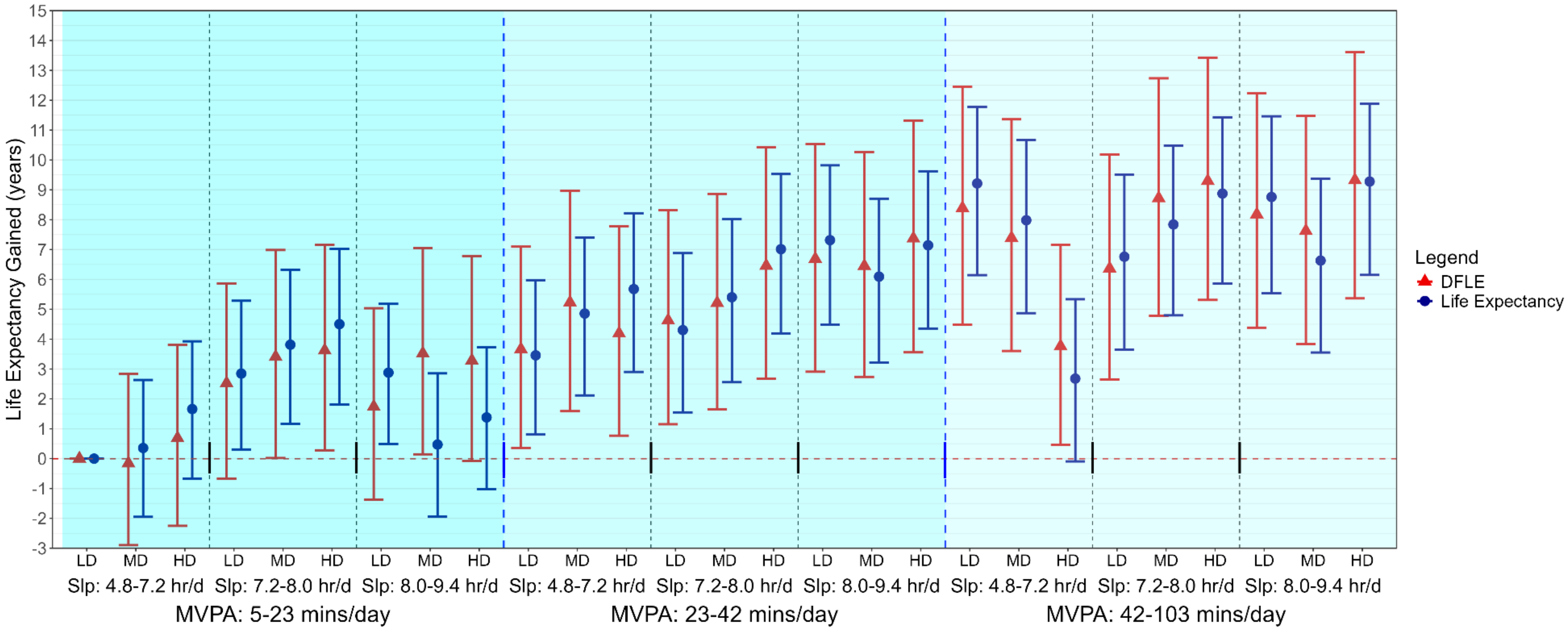


**Supplementary Figure 8:** Multivariable-adjusted associations of combined sleep, physical activity, and nutrition with lifespan and healthspan excluding individuals with an event in the first 3 years of follow-up (n = 58,610; events = 1,990)

**Legend**: Forest plot shows the SPAN associations with life expectancy (lifespan) and disease free life expectancy (DFLE; healthspan) after removing those with a mortality event in the first three years of follow-up. Model is adjusted for age, sex, ethnicity, smoking, education, Townsend deprivation index, alcohol, discretionary screen time (time spent watching TV or using the computer outside of work), light intensity physical activity, medication (blood pressure, insulin, and cholesterol), and familial history of CVD and cancer. Sleep (hours/day), physical activity (moderate to vigorous intensity (MVPA) minutes/day), and nutrition (Dietary Quality Score (DQS)) were included in the model as a joint term. The specific ranges for each exposure included sleep duration as 5.0-7.2 hours/day (low), 7.2-8.0 hours/day (medium), and 8.0-9.4 hours/day (high); MVPA measurements as 6-23 minutes/day (low), 23-42 minutes/day (medium), and 43-104 minutes/day (high); and diet quality using the DQS as 34.0-50.0 (low), 50.0-57.5 (medium), and 57.5-72.5 (high). The lowest tertiles for all three exposures (sleep, MVPA and DQS) was the referent group. Dashed blue lines separate tertiles MVPA and dashed black lines separate tertiles of sleep. Sleep (Slp); Low Diet Quality (LD); Medium Diet Quality (MD); High Diet Quality (HD).

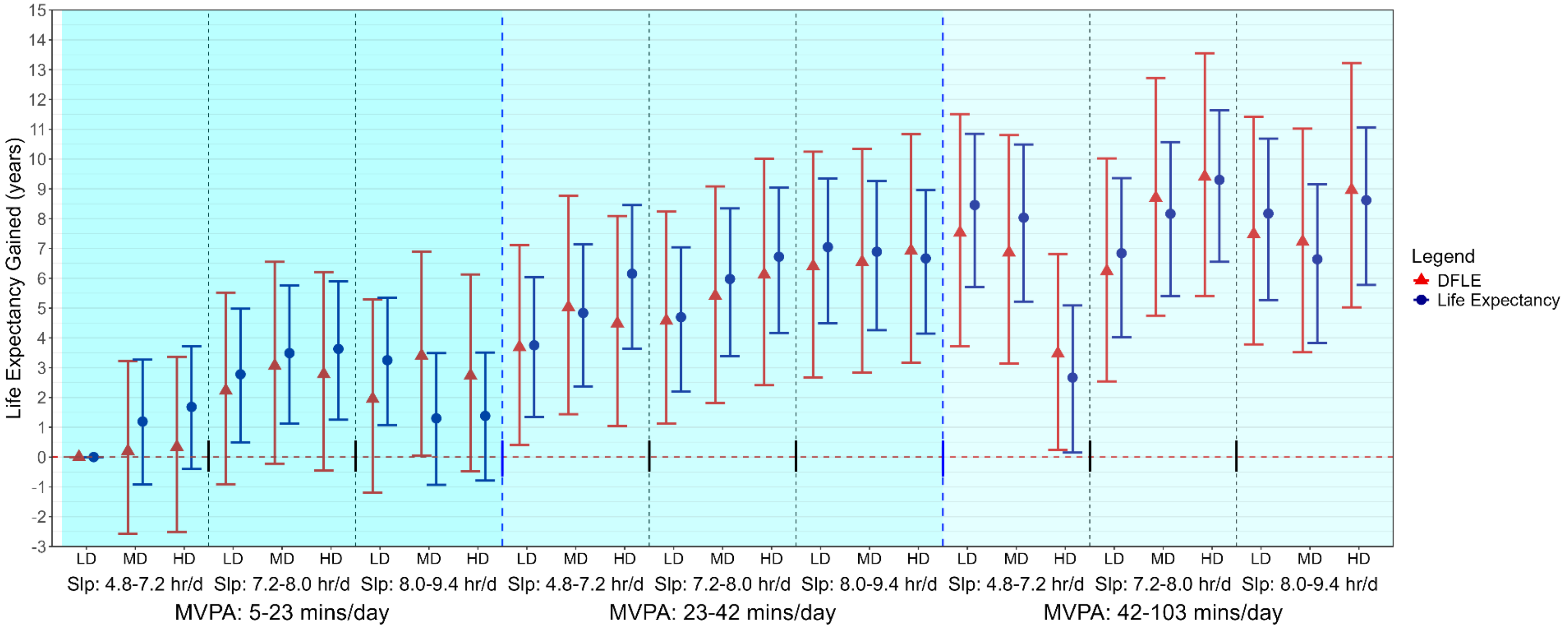


**Supplemental Figure 9**: Multivariable-adjusted associations of combined sleep, physical activity, and nutrition with lifespan and healthspan adjusted for BMI (n = 58,363; events = 2,405)

**Legend**: Life expectancy (lifespan) was estimated using life table models, with predictions based on hazard ratio-adjusted mortality rates for all-cause mortality associated with each category. Disease-free life expectancy (DFLE; healthspan) was calculated as the expected lifespan free from cardiovascular disease, cancer, type II diabetes, chronic obstructive pulmonary disease (COPD), or dementia. DFLE incorporated a life table approach that included age-specific incidence rates for each condition. Model is adjusted for age, sex, ethnicity, smoking, education, Townsend deprivation index, alcohol, discretionary screen time (time spent watching TV or using the computer outside of work), light intensity physical activity, medication (blood pressure, insulin, and cholesterol), previous diagnosis of major CVD (defined as disease of the circulatory system, arteries, and lymph, excluding hypertension), previous diagnosis of cancer, familial history of CVD and cancer, and BMI. Sleep (hours/day), physical activity (moderate to vigorous intensity (MVPA) minutes/day), and nutrition (Dietary Quality Score (DQS)) were included in the model as a joint term. The specific ranges for each exposure included sleep duration as 4.8-7.2 hours/day (low), 7.2-8.0 hours/day (medium), and 8.0-9.4 hours/day (high); MVPA measurements as 5-23 minutes/day (low), 23-42 minutes/day (medium), and 42-103 minutes/day (high); and diet quality using the DQS as 32.5-50.0 (low), 50.0-57.5 (medium), and 57.5-72.5 (high). The lowest tertiles for all three exposures (sleep, MVPA and DQS) were considered the reference group. Dashed blue lines separate tertiles MVPA and dashed black lines separate tertiles of sleep. Sleep (Slp); Low Diet Quality (LD); Medium Diet Quality (MD); High Diet Quality (HD).

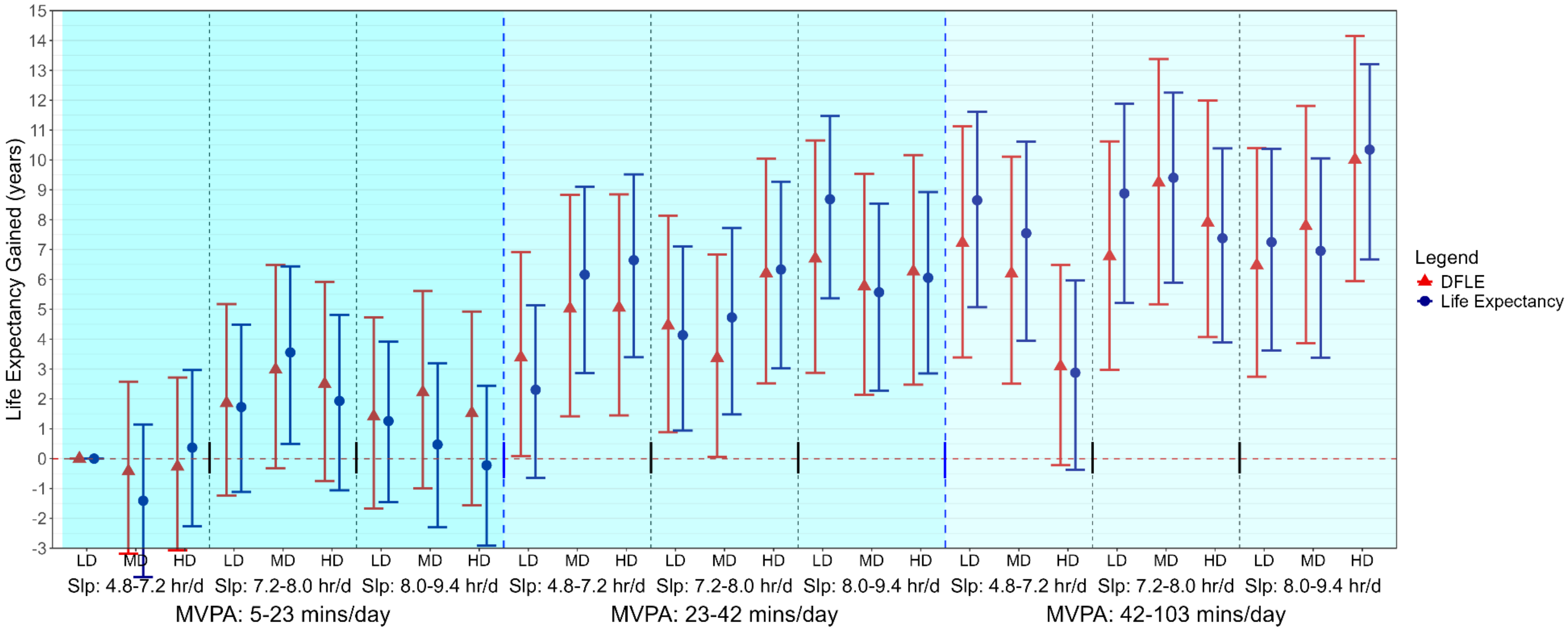


**Supplemental Figure 10:** Multivariable-adjusted associations of combined sleep, physical activity, and nutrition with lifespan and healthspan adjusted for sleep characteristics (n = 37,475; events = 1,506)

**Legend**: Life expectancy (lifespan) was estimated using life table models, with predictions based on hazard ratio-adjusted mortality rates for all-cause mortality associated with each category. Disease-free life expectancy (DFLE; healthspan) was calculated as the expected lifespan free from cardiovascular disease, cancer, type II diabetes, chronic obstructive pulmonary disease (COPD), or dementia. DFLE incorporated a life table approach that included age-specific incidence rates for each condition. Model is adjusted for age, sex, ethnicity, smoking, education, Townsend deprivation index, alcohol, discretionary screen time (time spent watching TV or using the computer outside of work), light intensity physical activity, medication (blood pressure, insulin, and cholesterol), previous diagnosis of major CVD (defined as disease of the circulatory system, arteries, and lymph, excluding hypertension), previous diagnosis of cancer, familial history of CVD and cancer, insomnia, snoring, chronotype (morning/evening person), and daytime sleepiness. Sleep (hours/day), physical activity (moderate to vigorous intensity (MVPA) minutes/day), and nutrition (Dietary Quality Score (DQS)) were included in the model as a joint term. The specific ranges for each exposure included sleep duration as 4.8-7.2 hours/day (low), 7.2-8.0 hours/day (medium), and 8.0-9.4 hours/day (high); MVPA measurements as 5-23 minutes/day (low), 23-42 minutes/day (medium), and 42-103 minutes/day (high); and diet quality using the DQS as 32.5-50.0 (low), 50.0-57.5 (medium), and 57.5-72.5 (high). The lowest tertiles for all three exposures (sleep, MVPA and DQS) were considered the reference group. Dashed blue lines separate tertiles MVPA and dashed black lines separate tertiles of sleep. Sleep (Slp); Low Diet Quality (LD); Medium Diet Quality (MD); High Diet Quality (HD).

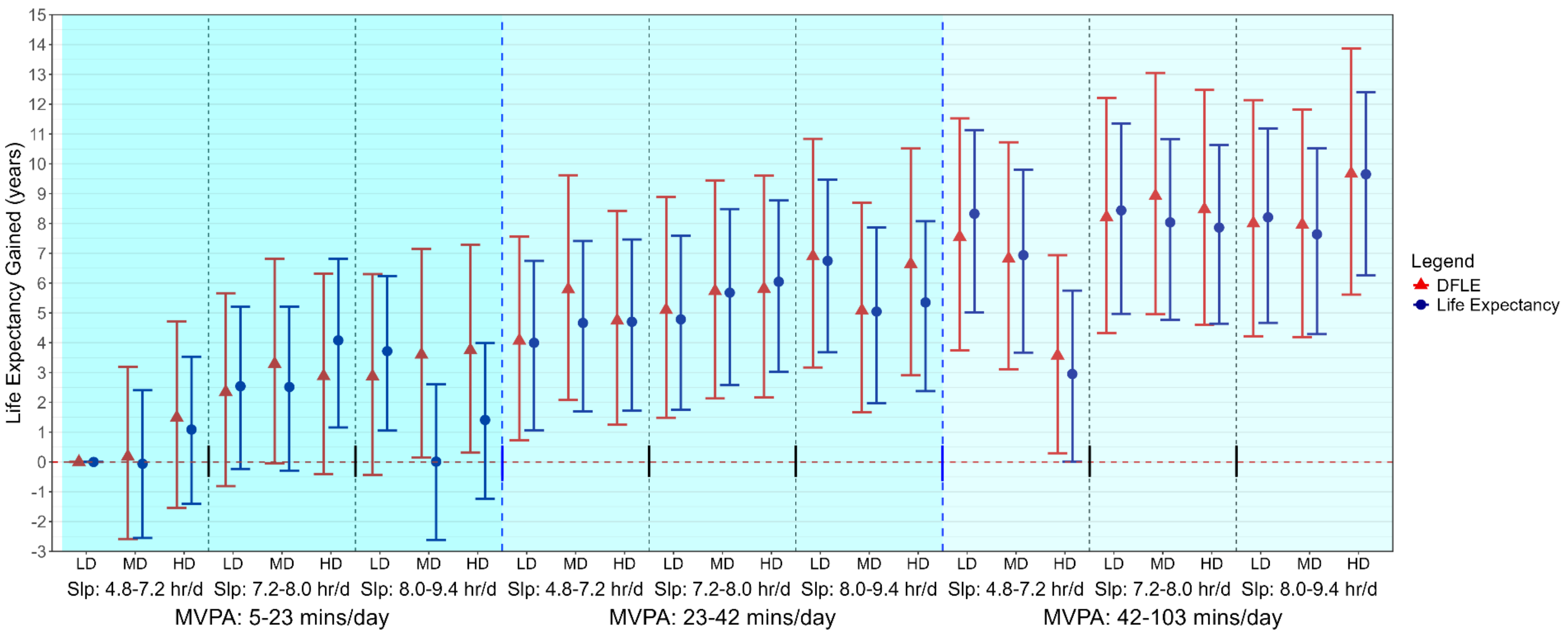


**Supplemental Figure 11:** Multivariable-adjusted associations of combined sleep, physical activity, and nutrition with adjusted lifespan and healthspan using the proportion of ultra-processed food (n = 43,694; events = 1,758)

**Legend**: Life expectancy (lifespan) was estimated using life table models, with predictions based on hazard ratio-adjusted mortality rates for all-cause mortality associated with each category. Disease-free life expectancy (DFLE; healthspan) was calculated as the expected lifespan free from cardiovascular disease, cancer, type II diabetes, chronic obstructive pulmonary disease (COPD), or dementia. DFLE incorporated a life table approach that included age-specific incidence rates for each condition. Model is adjusted for age, sex, ethnicity, smoking, education, Townsend deprivation index, alcohol, discretionary screen time (time spent watching TV or using the computer outside of work), light intensity physical activity, medication (blood pressure, insulin, and cholesterol), previous diagnosis of major CVD (defined as disease of the circulatory system, arteries, and lymph, excluding hypertension), previous diagnosis of cancer, familial history of CVD and cancer. From 2009-2012, dietary data was also collected using 1-4 separate 24-hour dietary recall for a subgroup of participants (n = 211,031)[1]. Diet quality was defined as the percentage of dietary ultra-processed food where higher diet quality had a lower proportion of ultra-processed food in the diet. Sleep (hours/day), physical activity (moderate to vigorous intensity (MVPA) minutes/day), and nutrition (ultra-processed food intake, % of total diet by weight) were included in the model as a joint term. The specific ranges for each exposure included sleep duration as 4.8-7.2 hours/day (low), 7.2-8.0 hours/day (medium), and 8.0-9.4 hours/day (high); MVPA measurements as 5-23 minutes/day (low), 23-42 minutes/day (medium), and 42-103 minutes/day (high); and diet quality using the proportion of ultra-processed food as 21.5-100.0 (low), 13.2-21.5 (medium), and 0.0-13.2% (high). The lowest tertiles for all three exposures (sleep, MVPA and DQS) were considered the reference group. Dashed blue lines separate tertiles MVPA and dashed black lines separate tertiles of sleep. Sleep (Slp); Low Diet Quality (LD); Medium Diet Quality (MD); High Diet Quality (HD).

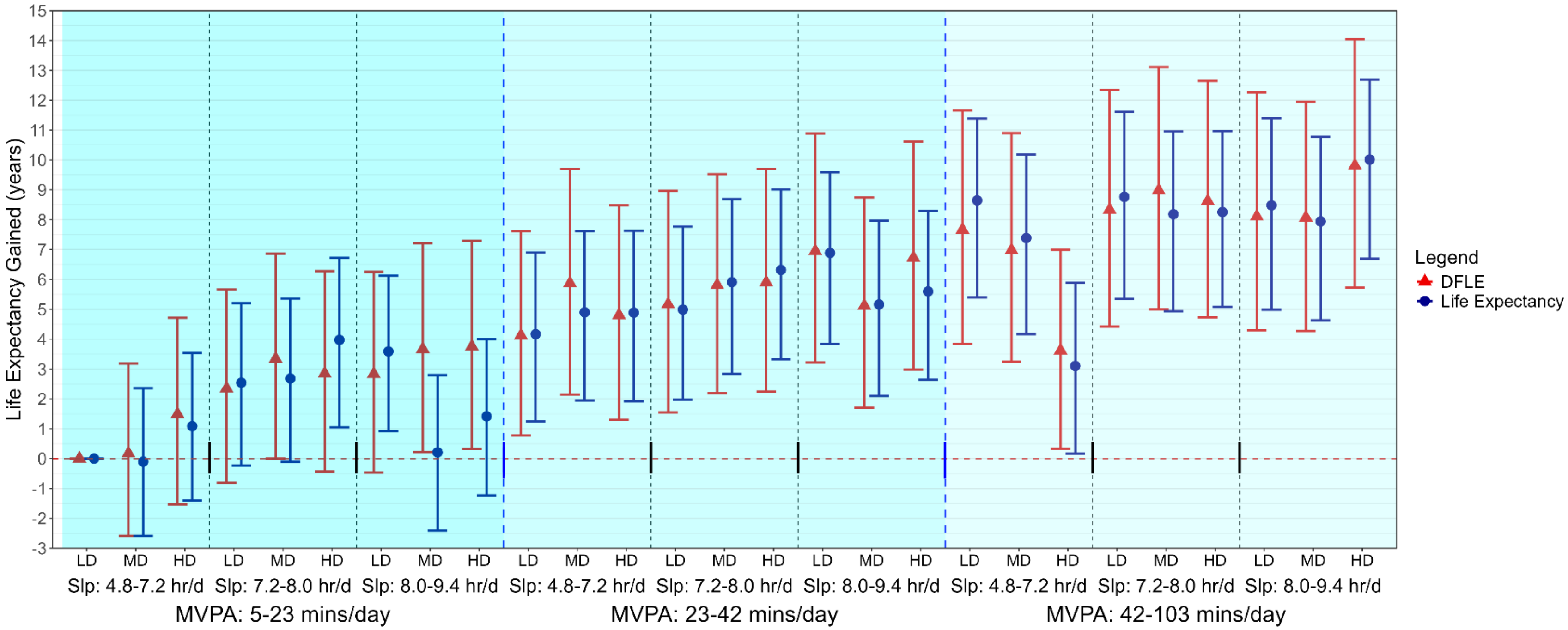


**Supplemental Figure 12:** Multivariable-adjusted associations of combined sleep, physical activity, and nutrition with lifespan and healthspan adjusted for total energy intake (n = 42,990; 1,758 events)

**Legend**: Life expectancy (lifespan) was estimated using life table models, with predictions based on hazard ratio-adjusted mortality rates for all-cause mortality associated with each category. Disease-free life expectancy (DFLE; healthspan) was calculated as the expected lifespan free from cardiovascular disease, cancer, type II diabetes, chronic obstructive pulmonary disease (COPD), or dementia. DFLE incorporated a life table approach that included age-specific incidence rates for each condition. Model is adjusted for age, sex, ethnicity, smoking, education, Townsend deprivation index, alcohol, discretionary screen time (time spent watching TV or using the computer outside of work), light intensity physical activity, medication (blood pressure, insulin, and cholesterol), previous diagnosis of major CVD (defined as disease of the circulatory system, arteries, and lymph, excluding hypertension), previous diagnosis of cancer, familial history of CVD and cancer, and total energy intake. From 2009-2012, dietary data was also collected using 1-4 separate 24-hour dietary recall for a subgroup of participants (n = 211,031)[1]. Energy intake outliers (<800 or >4200 kcal/ per day) for men and (<600 or >3500 kcal per day) for women were also excluded from this sample[2]. Sleep (hours/day), physical activity (moderate to vigorous intensity (MVPA) minutes/day), and nutrition (Dietary Quality Score (DQS)) were included in the model as a joint term. The specific ranges for each exposure included sleep duration as 4.8-7.2 hours/day (low), 7.2-8.0 hours/day (medium), and 8.0-9.4 hours/day (high); MVPA measurements as 5-23 minutes/day (low), 23-42 minutes/day (medium), and 42-103 minutes/day (high); and diet quality using the DQS as 32.5-50.0 (low), 50.0-57.5 (medium), and 57.5-72.5 (high). The lowest tertiles for all three exposures (sleep, MVPA and DQS) were considered the reference group. Dashed blue lines separate tertiles MVPA and dashed black lines separate tertiles of sleep. Sleep (Slp); Low Diet Quality (LD); Medium Diet Quality (MD); High Diet Quality (HD).

**Supplementary Methods 1:** Additional study design details

### Study Sample and Design

This study analyzed the data from the UK Biobank which is an ongoing prospective cohort study comprised of adults aged 40-69 at baseline (2006-2010)[3]. Participants provided informed consent and ethical approval was provided by the UK's National Health Service, National Research Ethics Service (Ref80 11/NW/0382). Deaths were ascertained through linkage with the National Health Service Digital of England and Wales or the National Health Service Central Register and National Records of Scotland. Censoring for England, Wales, and Scotland was up to November 30th, 2022.

Hospital inpatient data was ascertained through linkage with the National Health Service Digital for England, the Information and Statistics Division for Scotland, and Secure Anonymized Information Linkage for Wales. Censoring for England and Scotland was up to October 31st, 2022 and August 31st, 2022, respectively. Censoring for Wales was up to May 31st, 2022. Cancer data linkage was obtained through national cancer registries. For England and Wales, cancer diagnosis data were followed up through 31 December 2020 and 31 December 2016 respectively, and were provided by NHS England[4]. For Scotland, cancer diagnosis data were followed up through 30 November 2021 and provided by the National Records of Scotland[4].

Between 2013 and 2015 (median 5.5 years after the baseline measurements), 103,684 UK Biobank participants wore a wrist-worn accelerometer for 7 days[5]. The accelerometers were calibrated before being mailed to the individuals. During the initial processing stage, we excluded participants who if no sleep data was recorded, the accelerometer was poorly calibrated (>10 milli-gravitational units (mg)), or a faulty accelerometer was distributed (>100mg)[6-9]. We excluded participants with missing covariates and insufficient valid wear days. Monitoring days were considered valid if wear time was greater than 16 hours. To be included in the analysis, participants were required to have at least three valid monitoring days, with at least one of those days being a weekend day. We excluded participants who reported that they could not walk[6-9].

### Outcome ascertainment

Ascertainment of major chronic conditions

| Variable | Definition |
|---|---|
| Cardiovascular disease incidence[9] | CVD was defined as diseases of the circulatory system, excluding hypertension, diseases of arteries, and lymph. |

| | |
|---|---|
| | The ICD-10 codes included were: I0, I11, I13, I20-I51, I60-I69. |
| Cancer incidence definition[7] | The definition of total cancer excluded in situ, benign, uncertain, non-melanoma skin cancer, or non-well-defined cancers. The ICD-10 codes used were C15, C220, C221, C34, C649, C659, C160, C54, C559, C92, C900, C18, C260, C0, C11, C12, C13, C14, C30, C31, C32, C33, C34, C38, C390, C398, C399, C199, C209, C67, C50. |
| Type II diabetes[10] | Inpatient hospitalisation record ICD 10 codes: E11.<br><br>Read Codes using general practitioner records:<br><br><u>Version *2*:</u> C1041, C1096, C109A, C109B, C109C, C109E, C109F, C109G, C109H, C10F6, C10FA, C10FB, C10FC, C10FE, C10FF, C10FG, C10FH, C10FL, C10FM, C10FQ, C10FR.<br><br><u>Version *3*:</u> C1011, C102, C1021, C1031, C1041, C1051, C1061, C1071, C1074, C1090, C1091, C1092, C1093, C1094, C1095, C1096, C1097, C10y1, C10z1, X40J5, X40J6, X40JJ, Xaagf, XaCJ2, XaELQ, XaEnp, XaEnq, XaF05, XaFmA, XaFn7, XaFn8, XaFn9, XaFWI, XaIrf, XaIzQ, XaIzR, XaJQp, XaKyX, XE10F, XSETH. |
| Chronic Obstructive Pulmonary Disease[11] | Inpatient hospitalisation record ICD 10 codes:<br><br>.J41, J43, J44, J98.2, J98.3 |
| Dementia[12] | Inpatient hospitalisation record ICD 10 codes:<br>A81.0, F00, F00.0, F00.1, F00.2, F00.9, F01, F01.0, F01.1, F01.2, F01.3, F01.8, F01.9, F02, F02.0, F02.1, F02.2, F02.3, F02.4, F02.8, F03, F05.1, F10.6, G30, G30.0, G30.1, G30.8, G30.9, G31.0, G31.1, G31.8, I67.3<br><br>Read Codes using general practitioner records:<br><br><u>Version *2*:</u> 1461, A411., A4110, E00.., E000., E001., E0010, E0011, E0012, E0013, E001z, E002., E0020, E0021, E002z, E003., E004., E0040, E0041, E0042, E0043, E004z, E012., E02y1, E041., Eu00., Eu000, Eu001, Eu002, Eu00z, Eu01<br><br><u>Version *3*:</u> .1461, 1461, .E11., .E111, .E112, .E113, .E114, .E115, .E116, .E11Z, .F21Z, .F371, .G78., A411., |

| | |
|---|---|
| | A4110, E00.., E000., E001., E0010, E0011, E0012, E0013, E001z, E002., E0020, E0021, E002z, E003., E004., E0040, E0041 |

**Supplementary Methods 2**. Wearable behaviour classification methods

#### Sleep and non-wear time

The non-wear time was determined using a previously validated algorithm that uses wrist tilt angle to determine non-wear with 86-95% accuracy[13]. No values were imputed for non-wear time. Sleep was defined as the average daily duration of sleep (hours/day) as calculated using a validated algorithm based on relative changes in wrist tilt angle between successive 5-second windows[14]. For each interval of 5 seconds, the average of the estimated wrist tilt angle was calculated and a rolling 5-minute median served as an input for the algorithm to identify sleep onset and sleep offset, and then time spent asleep within this timeframe[13,14].

#### Two-stage random forest physical activity intensity and posture classification

Incidental physical activity was classified using a validated two-stage random forest activity classifier that first classifies each 10 second window (epoch) as sedentary (lying or sitting still), stationary plus (active sitting, standing still, active standing), walking, or running (**Diagram A**)[13-15]. These activities were then classified into one of four activities including: sedentary, light, moderate, and vigorous. Walking activities (gardening, active commuting, etc) were classified by normalized gravitational units (g) where <100 milli g were classified as light intensity (<3 METs), ≥100 milli g and <400 milli g were considered moderate intensity physical activity (≥3 to <6 METs), and ≥400 milli g were considered vigorous-intensity PA (≥6 METs)[15]. All windows classified as running/high energetic activity were classified as vigorous-intensity physical activity (≥ 6 METs)[6,8,15]. A major advantage of this classification approach is the lower risk of possible misclassification of sporadic high-accelerations that may occur during certain stationary light activities (e.g., dishwashing)[16-18].

**Physical Activity Classification Scheme (Diagram A)**

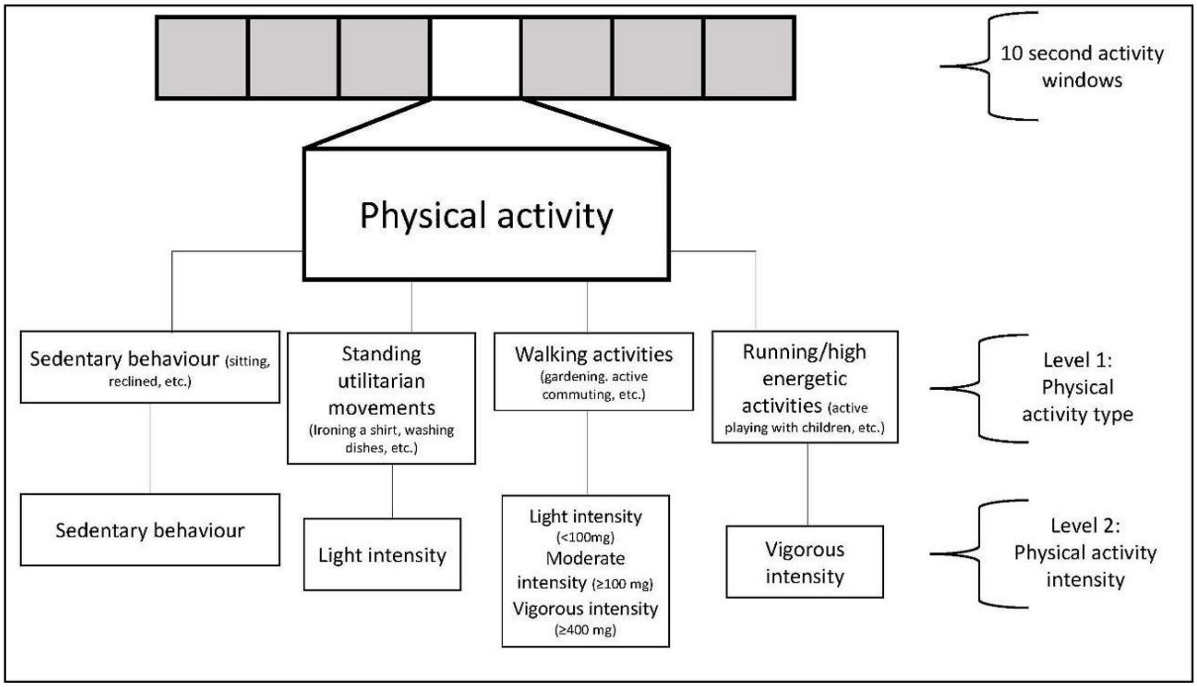


**Physical Activity Classification Performance**

The performance of this physical activity classification scheme was tested in an independent sample of 102 adults from the US[19] and Australia[20]. This data includes direct observation measurement of 105,767 activity samples from structured and free-living activities (17,627 minutes), which were used to test the robustness and generalisability of the two-stage activity and intensity classifier. The data was collected from participant-worn or researcher-held Go-Pro video recordings. All data was imported into Noldus Observer XT software for continuous video coding. The direct observation coding generated continuous physical activity codes corresponding to the start and finish of each movement. These coded movements were then compared against the accelerometer data using the available time-stamp information. The table below includes the performance metrics across activities. Interobserver reliability was assessed by dual coding. The intraclass correlation coefficient for coding activities was 0.912 (0.866-0.942). The performance in metrics and confusion matrix for activity classification is shown below.

**Classifier Performance Metrics for Intensity in US and Australian Adults**

| | Sensitivity | Specificity | Precision | F-score | Overall Accuracy | Weighted Kappa | Overall F-score |
|---|---|---|---|---|---|---|---|
| Sedentary | 86.5 | 93.7 | 90.5 | 88.5 | | | |
| Light | 71.2 | 89.4 | 55.8 | 62.6 | | | |
| Moderate | 85.4 | 96.6 | 92.7 | 88.9 | | | |
| Vigorous | 95.4 | 99.4 | 94.6 | 95.0 | | | |

| | | | | | | | |
|---|---|---|---|---|---|---|---|
| | | | | | **84.6** | 0.78 | 83.8 |

Rows= ground truth; columns=predictions; bold=correct classification; all activities were free-living or simulated free-living activities.

**Confusion Matrix for Activity Classification in US and Australian Adults**

| | Sedentary | Light | Moderate | Vigorous |
|---|---|---|---|---|
| Sedentary | **36,904** | 5,232 | 508 | 2 |
| Light | 3,120 | **11,712** | 1,612 | 17 |
| Moderate | 502 | 4,016 | **29,528** | 526 |
| Vigorous | 226 | 17 | 214 | **9,470** |

Rows= ground truth; columns=predictions; bold=correct classification; all activities were free-living or simulated free-living activities.

**Supplementary Methods 3**. Calculation of lifespan and healthspan

Life expectancy (lifespan) and disease-free life expectancy (healthspan) were calculated using a traditional life table approach[21-24] which use three pieces of information including:

1) All-cause mortality risk for lifestyle combinations derived from multivariable adjusted all-cause mortality model using a sample of adults from the UK Biobank wearables sub-study[25]
2) Age and sex-specific mortality rates estimated from UK Census Data[21]
3) Age-specific prevalence and incidence rates of the five leading contributors of chronic disease including cardiovascular disease, cancer, type II diabetes, chronic obstructive pulmonary disease, and dementia

Life tables can be conducting the below functions at each age in the table:

$l_x$: The number of people alive at the start of age $x$

$q_x$: The probability of dying between ages $x$ and $x$ +1

$d_x$: The number of deaths between ages $x$ and $x$ + 1, calculated as

$$d_x = l_x * q_x$$

$L_x$: The person-years lived between ages $x$ and $x$ +1 approximated as

$$L_x = l_x - \frac{d_x}{2}$$

$T_x$: The total person-years lived above age $x$, calculated as

$$T_x = \sum_x L_x$$

$e_x$: Life expectancy at age $x$, calculated:

$$e_x = \frac{T_x}{l_x}$$

In this study, life tables were constructed for the total sample and sex-specific groups, incorporating female- and male-specific mortality rates proportionally in the dataset and sex-stratified analyses. Mortality rates were extracted from UK Census data[21] across sex-specific 5-year age bands. In the present study, life expectancy at age 40 and ending at age 100 was used, aligning with the age of the core analytical sample. As previously described[26], we then fitted a multi-variable all-cause mortality model for the joint tertile-based sleep, physical activity, and nutrition (SPAN) exposure to extract hazard ratios (HR) for each mutually exclusive category. Changes in life expectancy associated with varying levels of sleep, physical activity, and nutrition were calculated by incorporating the derived HRs into the life table to reflect the change in mortality risk[22,27]. To estimate the gain in life expectancy, the predicted life expectancy for the referent group was then subtracted from the life expectancy of each SPAN combination. The same methodology was used to estimate life expectancy with the individual behaviours and the continuous SPAN score which combined these behaviours with equal weight on a scale from 0 to 100.

Disease-free life expectancy was calculated as an extension of the life table approach[21,22], accounting for the age-specific prevalence and incidence of the five leading contributors to disease burden in the UK[28], including cardiovascular disease, type II diabetes, cancer, and chronic obstructive pulmonary disease. The incidence in each stratified SPAN group was estimated to assess the burden of disease at each age, assuming static rates with no dynamic changes over time.

**Disease-Free Life Expectancy (healthspan): Continuation of the Life Table**

Partition years lived ($L_x$) into $L_x^{Healthy}$ and $L_x^{Diseased}$ by considering the prevalence and incidence of disease $P_x$ at each age ($x$):

Calculate healthy person-years at age $x$:

$$L_x^{Healthy} = L_x * (1 - P_x)$$

Calculate the number of diseased years at age $x$

$$L_x^{Diseased} = L_x * P_x$$

Sum the years of healthy living:

$$T_x^{Healthy} = \sum_x L_x^{Healthy}$$

Calculate disease-free life expectancy

$$e_x^{Healthy} = \frac{T_x^{Healthy}}{l_x}$$

The calculation was then performed within the joint-tertile-based SPAN analyses, where individual life tables were created for each mutually exclusive category, incorporating shifts in disease prevalence. The disease-free life expectancy was then calculated as an extension of the primary life table, partitioning the years lived at each age into those with and without chronic conditions to estimate the total years lived free of disease. To address variability and uncertainty in disease incidence and prevalence across groups, we used a Monte Carlo simulation with 10,000 iterations to estimate disease-free life expectancy[29-31]. To estimate the minimum change for disease-free life expectancy, the SPAN score was transformed back into combined SPAN components using a previously established relative contribution from each behaviour in their combined association with mortality[26].

**Supplementary Methods 4**. Calculation of the composite SPAN score

To explore the minimum effective behaviour changes[26] needed for meaningful improvements in lifespan and healthspan, we constructed a continuous composite SPAN score scored equally (i.e., 33.3 points per behaviour) according to the dose-response relationship of each behaviour with mortality. The scoring of each exposure within the composite SPAN score was determined based on the theoretically optimal levels identified from the dose-response relationship with all-cause mortality[32,33].

For example, we identified a U-shaped dose-response relationship between sleep and all-cause mortality, with the lowest risk of mortality observed at 7.9 hours per day of sleep. To account for this in the score, the theoretically most optimal sleep duration of 7.9 hours would be scored 33.3 points, while sleep less or more than this amount would be scored proportionately as it deviates from this point. For physical activity and diet, the relationship was positively linear with all-cause mortality and was scored proportionally from 0-33.3 based on their relative rank according to the minimum and maximum of each behaviour. The final composite SPAN score was the sum of these three behaviour scores, ranging from 0-100, where higher values indicate theoretically healthier.

**The formula is described as:**

$$\text{SPAN\_score} = \frac{\text{Physical\_Activity} - \min(\text{Physical\_Activity})}{\max(\text{Physical\_Activity}) - \min(\text{Physical\_Activity})} \times 33.33 + \frac{\text{Diet} - \min(\text{Diet})}{\max(\text{Diet}) - \min(\text{Diet})} \times 33.33 + \left[33.33 - \frac{|\text{Sleep} - 7.9|}{\max(|\text{Sleep} - 7.9|)} \times 33.33\right]$$

**Supplementary Table 1.** Diet quality score index for food-frequency questionnaire dietary data

| Food components | UK Biobank field ID | Amount per serving | Criteria for maximum score (10) | Criteria for minimum score (0) |
|---|---|---|---|---|
| Fruit | 1309 (pieces fresh fruit/day) 1319 (pieces dried fruit/day) | 1309 – 1 piece<br>1319 – 5 pieces | ≥3 servings/day | 0 servings/day |
| Vegetable | 1289 (tablespoons cooked vegetables/day)<br>1299 (salad/raw vegetables/day) | 3 heaped tablespoons | ≥3 servings/day | 0 servings/day |
| Whole grains | 1438, 1448 (wholemeal/wholegrain bread slices/week)<br>1458, 1468 (bran/oat/muesli cereal) | 1438/1448 – 1 slice/day<br>1458/1468 – 1 bowl/day | ≥3 servings/day | 0 servings/day |
| Fish | 1329 (oily fish/week)<br>1339 (non-oily fish/week) | Once/week | ≥2 servings/week | 0 servings/week |
| Dairy | 1408 (cheese/week)<br>1418 (milk type) | 1408 – 1 piece/day<br>1418 – 1 glass/day if consumption of any type of milk | ≥2 servings/day | 0 servings/day |
| Vegetable oils | 1428 (Flora Pro-Active/Benecol spread)<br>2654 (Flora Pro-Active/Benecol, soft margarine -, olive oil based -, polyunsaturated/sunflower oil based -, other low/reduced fat spread)<br>1438 (bread slices/week) | 1 serving/day if in combination with eating at least 2 slices of bread (ID 1438) | ≥2 servings/day | 0 servings/day |
| Refined grains | 1438, 1448 (white, brown, other bread slices/week) 1458, 1468 (biscuit, other cereals/week) | 1438/1448 – 1 slice/day<br>1458/1468 – 1 bowl/day | 0 servings/day | >2 servings/day |
| Processed meats | 1349 (processed meat/week or daily)<br>3680 (age when last ate meat) | 1349 – 1 piece/day<br>3680 – 0 pieces/day if indicated having never eaten meat | 0 serving/week | >1 serving/week |
| Unprocessed red meats | 1369 (beef/week or day)<br>1379 (lamb or mutton/week or day)<br>1389 (pork/week or day)<br>3680 (age when last ate meat) | 1359-1389 – once/week<br>3680 – 0 pieces/day if indicated having never eaten meat | 0 serving/week | >2 serving/week |
| Sugar-sweetened beverages | 6144 (never consumes drinks containing sugar) | 0 servings | Don't drink | Drink |

Diet quality score information is adapted from previously established work by Zhuang *et al*. Diabetes Care[34]. Intermediate intake for each dietary component were scored relative to minimum to maximum intake of each food component. The formula for intermediate intakes of adequacy components is described as: component score = (maximum score / (Amax - Amin))*(X - Amin) and for moderate components (refined grains, processed meat, and unprocessed red meat) component score = (maximum score - maximum score / (Amax - Amin))*(X - Amin). The food frequency questionnaire demonstrated moderate reproducibility for food groups (Intraclass Correlation Coefficient (ICC): 0.48-0.66) and modest agreement with alternative dietary intake measures from the 24-hour recall (ICC: 0.38-0.63). This level of agreement and reproducibility is comparable to previous prospective observational studies[35-37]. The food frequency questionnaire has also been validated against the 24-hour dietary recall using objective biomarkers as the standard[38].

**Supplementary Table 2:** Mortality and disease events across the mutually exclusive sleep, physical activity, and nutrition combinations

| | | Nutrition Low | Nutrition Medium | Nutrition High |
|---|---|---|---|---|
| **MVPA Low** | **Sleep Low** | n = 2542 (Events = 203) | n = 2234 (Events = 163) | n = 2205 (Events = 166) |
| | **Sleep Medium** | n = 2178 (Events = 116) | n = 1982 (Events = 100) | n = 1861 (Events = 100) |
| | **Sleep High** | n = 2458 (Events = 138) | n = 2136 (Events = 138) | n = 2097 (Events = 152) |
| **MVPA Medium** | **Sleep Low** | n = 2312 (Events = 98) | n = 2116 (Events = 83) | n = 1945 (Events = 77) |
| | **Sleep Medium** | n = 2298 (Events = 85) | n = 2241 (Events = 75) | n = 2092 (Events = 69) |
| | **Sleep High** | n = 2339 (Events = 67) | n = 2180 (Events = 66) | n = 2169 (Events = 73) |
| **MVPA High** | **Sleep Low** | n = 2271 (Events = 48) | n = 2062 (Events = 50) | n = 2006 (Events = 92) |
| | **Sleep Medium** | n = 2407 (Events = 55) | n = 2386 (Events = 52) | n = 2247 (Events = 46) |
| | **Sleep High** | n = 2113 (Events = 46) | n = 2127 (Events = 56) | n = 2074 (Events = 44) |

The sample size and number of all-cause mortality events for each Sleep, Physical Activity, and Nutrition group is detailed above (n = 59,078; events = 2,458). Participants were grouped by Sleep, Physical Activity, and Nutrition exposure tertiles (i.e., low, moderate, and high) which equated to a joint exposure of 27 separate groups for all three behaviours. The specific ranges for each exposure included sleep duration as 4.8-7.2 hours/day (low), 7.2-8.0 hours/day (medium), and 8.0-9.4 hours/day (high); moderate to vigorous physical activity (MVPA) measurements as 5-23 minutes/day (low), 23-42 minutes/day (medium), and 42-103 minutes/day (high); and diet quality using the DQS as 32.5-50.0 (low), 50.0-57.5 (medium), and 57.5-72.5 (high).

**Supplementary Table 3.** Covariate Definitions.

| Variable | Definition | UK Biobank field ID (if applicable) |
|---|---|---|
| Age | Categorical (4) | 34, 52, accelerometer date-timestamp |
| Sex | Female/Male | 31 |
| Ethnicity | White/Others | 21000 |
| Education | College/University; A/AS level; O levels; CSE; NVQ/HND/HNC; other | 6138 |
| Smoking status | Never, past, current | 20116 |
| Alcohol consumption | Units/day | 20403 |
| Light intensity physical activity | Standing utilitarian movements, slow walking (<3 METs) | Derived from accelerometer data |
| Discretionary screen-time | Self-reported time spent/day watching TV and using a computer outside of work | 1070, 1080 |
| Townsend deprivation | Categorical (5) | 22189 |
| Use of cholesterol medication | Yes/No | 6177, 6153 |
| Use of blood pressure medication | Yes/No | 6177, 6153 |
| Use of diabetes medication | Yes/No | 6177, 6153 |
| Previous CVD | Identified by self-report and hospitalisation. Defined as disease of the circulatory system, arteries, and lymph, excluding hypertension | 20002, 41270 |
| Previous cancer | Identified by self-report and cancer registry. | 20001, 100092 |
| Familial history of CVD | Self-reporter mother of father diagnosed with heart disease or stroke | 20107, 20110 |
| Familial history of cancer | Self-reporter mother of father diagnosed with cancer | 20107, 20110 |
| High frailty scale | Categorical (yes/no); high frailty indicates a score of ≥3 on a 0 to 5 | 2306, 120107, 2624, 1011, 3637, 991, 971, 924, 46, 47 |
| Body mass index | Continuous; kilogram/meter$^2$ | 23104 |
| Total energy intake | Continuous, kcal/day derived from 24-hour dietary recall data | 26002 |
| Morning/evening person (chronotype) | Categorical (definitely a ‘morning’ person; more a ‘morning’ person than ‘evening’ person; more an ‘evening’ person than ‘morning’ person; definitely an ‘evening’ person) | 1180 |

| | | |
|---|---|---|
| Insomnia | Categorical (never/rarely; sometimes; usually) | 1200 |
| Snoring | Categorical (yes/no) | 1210 |
| Daytime sleepiness | Categorical (never/rarely; sometimes; often) | 1220 |

Additional detail is available online at https://biobank.ndph.ox.ac.uk/showcase/.

**Supplementary Table 4:** Model variance inflation factors for combined SPAN behaviours

| Primary Model with Combined SPAN Behaviours | |
|---|---|
| **Variable** | **Variance inflation factor (VIF)** |
| Sleep, moderate to vigorous physical activity, and nutrition (combined variable) | 1.28 |
| Age (self-report) | 1.17 |
| Sex (self-report) | 1.17 |
| Ethnicity (self-report) | 1.03 |
| Smoking (self-report) | 1.10 |
| Alcohol (self-report) | 1.15 |
| Education (self-report) | 1.08 |
| Socioeconomic status (self-report) | 1.06 |
| Light physical activity (accelerometry derived) | 1.14 |
| Previous CVD (self-report) | 1.15 |
| Previous cancer (self-report) | 1.03 |
| Familial history of CVD (self-report) | 1.01 |
| Familial history of cancer (self-report) | 1.01 |
| Discretionary screen time (self-report) | 1.09 |
| Medication (self-report) | 1.21 |

The table provides the variance inflation factor (VIF) for each covariate in the primary analytical model (combined SPAN behaviours) adjusted for self-reported discretionary screen time and the sensitivity model adjusted for accelerometry derived sedentary behaviour. VIF values measure multicollinearity among the exposure variables, with a value of 1 indicating no correlation with other predictors. Higher values suggest increasing multicollinearity, with values greater than 5 indicating potentially problematic multicollinearity[39].

**Supplementary Table 5.** NOVA classification of food groups for 24-hour dietary recall data.

| NOVA classification level | UK Biobank field ID (if applicable) |
|---|---|
| Level four | Added sugars and preserves (26064), Animal fat spread lower fat (26062), Animal fat spread normal (26063), Biscuit cereal (26075), Biscuits (26068), Bran cereal (26076), Breaded/battered chicken (26069), Breaded/battered fish (26070), Chocolate confectionery (26080), Cream (26154), Fried/roast potatoes (26119), Low/non sugar sugar-sweetened beverages (26126), Mashed potatoes (26120), Meat substitutes - soy (26137), Meat substitutes - vegetarian (26145), Milk-based and powdered drinks (26087), Milk-dairy desserts (26084), Mixed bread brown and seeded (26071), Muesli (26105), Nut-based spreads (26106), Other cereal (sugar) (26079), Other desserts and cakes and pastries (26085), Other sweets (26140), Pizza (26116), Plant-based spread lower fat (26111), Plant-based spread normal (26112), Processed meat (26122), Samosa, pakora (26128), Sauces and condiments (high fat) (26129), Sauces and condiments (low fat) (26130), Savoury crackers (26083), Savoury snacks (26134), Soy desserts and yogurt (26086), Sugar-sweetened beverages and other sugary drinks (26127), Sushi (26139), Vegetable dips (26144) |
| Level three | High fat cheese (26099), Medium and low fat cheese (26103), White fish and tinned tuna (26149), White bread (26073), Wholemeal bread (26074), Other bread (26072) |
| Level two | Grain dishes - added fat (26097), Olive oil (drizzling/dunking) (26110) |
| Level one | Allium vegetables (26065), Apples and pears (26089), Beef (26066), Berries (26090), Citrus (26091), Coffee, caffeinated (26081), Coffee, decaffeinated (26082), Dried fruit (26092), Egg and egg dishes (26088), Fruit juice (26095), Green leafy/cabbages (26098), Lamb (26100), Legumes and pulses (26101), Oat cereal (non sugar) (26077), Oat cereal (sugar) (26078), Low fat yogurt (26102), Full fat yogurt (26096), Oily fish (26109), Other fruit (26093), Other meat, offal (26104), Other vegetables, including mushrooms, fruiting and mixed vegetables (26146), Peas and sweetcorn (26115), Pork (26117), Potatoes and sweet potatoes (baked/boiled) (26118), Poultry (26121), Raw salad (26123), Root vegetables (26125), Salted nuts and seeds (26108), Semi skimmed milk (26131), Rice/oat milk (26124), Shellfish (26132), Skimmed milk and cholesterol-lowering milk (26133), Soups (26135), Soy milk (26136), Stewed fruit (26094), Tea (26141), Tea, decaffeinated (26142), Tomatoes (26143), Unsalted nuts and seeds (26107), White pasta and rice (26113), Whole milk (26150), Wholemeal pasta, brown rice and other wholegrains (26114) |

From 2009-2012, dietary data was also collected using 1-4 separate 24-hour dietary recall for a subgroup of participants (n = 211,031)[2]. Additional detail on reproducibility and agreement between FFQ and the 24-hour dietary recall has been published elsewhere[40,41]. Food groups in each NOVA classification level were reported as the average weight (gram/day) from the 24-hour dietary recalls. Ultra-processed food intake was defined as the percentage of level four NOVA food groups relative to the average reported total food weight. All food categories and the definition of ultra-processed food intake were determined using a previously established method[42,43].

### **Supplementary Table 6.** STROBE.

| | Item No | Recommendation | Page No |
|---|---|---|---|
| **Title and abstract** | 1 | (*a*) Indicate the study's design with a commonly used term in the title or the abstract | |
| | | (*b*) Provide in the abstract an informative and balanced summary of what was done and what was found | 1-2 |
| **Introduction** | | | |
| Background/rationale | 2 | Explain the scientific background and rationale for the investigation being reported | 4-5 |
| Objectives | 3 | State specific objectives, including any prespecified hypotheses | 5 |
| **Methods** | | | |
| Study design | 4 | Present key elements of study design early in the paper | 6 |
| Setting | 5 | Describe the setting, locations, and relevant dates, including periods of recruitment, exposure, follow-up, and data collection | 6 |
| Participants | 6 | (*a*) Give the eligibility criteria, and the sources and methods of selection of participants. Describe methods of follow-up | 6 |
| | | (*b*) For matched studies, give matching criteria and number of exposed and unexposed | NA |
| Variables | 7 | Clearly define all outcomes, exposures, predictors, potential confounders, and effect modifiers. Give diagnostic criteria, if applicable | 6-7 |
| Data sources/ measurement | 8* | For each variable of interest, give sources of data and details of methods of assessment (measurement). Describe comparability of assessment methods if there is more than one group | 6-7 |
| Bias | 9 | Describe any efforts to address potential sources of bias | 8 |
| Study size | 10 | Explain how the study size was arrived at | Supplementary Figure 1 |
| Quantitative variables | 11 | Explain how quantitative variables were handled in the analyses. If applicable, describe which groupings were chosen and why | 6-7 |
| Statistical methods | 12 | (*a*) Describe all statistical methods, including those used to control for confounding | |
| | | (*b*) Describe any methods used to examine subgroups and interactions | 7-10 |
| | | (*c*) Explain how missing data were addressed | |
| | | (*d*) If applicable, explain how loss to follow-up was addressed | |
| | | (*e*) Describe any sensitivity analyses | |
| **Results** | | | |
| Participants | 13* | (a) Report numbers of individuals at each stage of study—eg numbers potentially eligible, examined for eligibility, confirmed eligible, included in the study, completing follow-up, and analysed | Supplemental figure 1 |
| | | (b) Give reasons for non-participation at each stage | Supplemental figure 1 |
| | | (c) Consider use of a flow diagram | Supplemental figure 1 |

| | | | |
|---|---|---|---|
| Descriptive data | 14* | (a) Give characteristics of study participants (eg demographic, clinical, social) and information on exposures and potential confounders | Table 1 |
| | | (b) Indicate number of participants with missing data for each variable of interest | Supplemental figure 1 |
| | | (c) Summarise follow-up time (eg, average and total amount) | Table 1 |
| Outcome data | 15* | Report numbers of outcome events or summary measures over time | |
| Main results | 16 | (*a*) Give unadjusted estimates and, if applicable, confounder-adjusted estimates and their precision (eg, 95% confidence interval). Make clear which confounders were adjusted for and why they were included | 10-14 |
| | | (*b*) Report category boundaries when continuous variables were categorized | Figure 1 Legend |
| | | (*c*) If relevant, consider translating estimates of relative risk into absolute risk for a meaningful time period | NA |
| Other analyses | 17 | Report other analyses done—eg analyses of subgroups and interactions, and sensitivity analyses | 14 |
| **Discussion** | | | |
| Key results | 18 | Summarise key results with reference to study objectives | 15 |
| Limitations | 19 | Discuss limitations of the study, taking into account sources of potential bias or imprecision. Discuss both direction and magnitude of any potential bias | 17-18 |
| Interpretation | 20 | Give a cautious overall interpretation of results considering objectives, limitations, multiplicity of analyses, results from similar studies, and other relevant evidence | 15-18 |
| Generalisability | 21 | Discuss the generalisability (external validity) of the study results | 17-19 |
| **Other information** | | | |
| Funding | 22 | Give the source of funding and the role of the funders for the present study and, if applicable, for the original study on which the present article is based | 20 |

*Give information separately for exposed and unexposed groups.

**Note:** An Explanation and Elaboration article discusses each checklist item and gives methodological background and published examples of transparent reporting. The STROBE checklist is best used in conjunction with this article (freely available on the Web sites of PLoS Medicine at http://www.plosmedicine.org/, Annals of Internal Medicine at http://www.annals.org/, and Epidemiology at http://www.epidem.com/). Information on the STROBE Initiative is available at http://www.strobe-statement.org.
